# Cross-Species and Tissue-Agnostic Prediction of Human Cancer Treatment Response using AI-Powered Cellular Morphometric Biomarkers from a Genetically Diverse Erbb2 Mouse Model

**DOI:** 10.1101/2025.09.15.25335730

**Authors:** Manuel J Pérez-Baena, Adrián Blanco-Gómez, Alejandro Jiménez-Navas, Marta Rodríguez-González, María del Mar Abad-Hernández, Mark A Taylor, Marina Mendiburu-Eliçabe, Natalia García-Sancha, Roberto Corchado-Cobos, César Augusto Rodríguez-Sánchez, Juan Jesús Cruz Hernández, Emilio Fonseca, Kuang-Yu Jen, Marina Holgado-Madruga, Rosemary J. Akhurst, Allan Balmain, Jian-Hua Mao, Jesús Pérez-Losada, Hang Chang

## Abstract

The success of drug development relies heavily on the use of animal models. However, increasing evidence shows that discoveries in these models often fail to translate to human patients. In this study, we developed an AI framework to discovery cross-species and tissue-agnostic cellular morphometric biomarkers (CMBs) as a new avenue to improve translatability. Using this framework, we identified CMBs and CMB tumor subtypes from treatment-naïve needle biopsies of mammary tumors in a genetically diverse *Erbb2/Neu* mouse model, showing significant correlation with responses to docetaxel treatment. These CMBs were then successfully translated to human patients with breast cancer, ovarian cancer or lung cancer, showing superior predictive/prognostic power to biomarkers and/or machine learning systems specifically optimized in human patients. Furthermore, Co-enrichment analysis revealed significantly conserved biological functions associated with CMBs in both mice and humans, such as cell cycle regulation, underscoring their relevance to tumor biology, treatment response and translatability.

## INTRODUCTION

Cancer exhibits significant heterogeneity, characterized by diverse morphological, genetic, and epigenetic features, resulting in varied clinical outcomes and treatment responses (1,2). This heterogeneity complicates the development of effective treatment strategies and underscores the need for precision medicine (3,4). Identifying reliable biomarkers that predict treatment responses and clinical outcomes is essential for advancing personalized cancer therapies (5).

Advancements in artificial intelligence (AI) and machine learning (ML) have introduced novel approaches for cancer classification and prognosis prediction (6,7). Cellular morphometric biomarkers (CMBs) are identified by AI/ML through the analysis of whole slide images (WSIs) of histologically stained tissue sections, which have emerged as promising tools for both precision oncology (8–10) and comparative oncology (11). CMBs are quantifiable features that capture key cellular and tissue morphology aspects, such as shape, size, texture, and spatial organization. They provide detailed insights into the tumor and its microenvironment, which traditional molecular biomarkers may overlook. Recently, the CMB-ML pipeline has been developed to allow the efficient and effective mining of robust and interpretable CMBs and CMB tumor subtypes with extensively validated translational and clinical value (8–13).

The success of developing drugs and other therapeutic approaches relies heavily on animal models in preclinical studies. The rigor and reproducibility of animal models are recognized as key factors facilitating the translation of findings in animal studies to successful clinical trials. Translating CMBs across species allows us to understand human disease evolution better, determine to what extent animal model systems reflect human biology, and facilitate the development of better drugs and more accurate predictions of toxicity. Therefore, CMBs likely lead to major advances in the translatability of animal models. We validated this possibility utilizing MMTV-*Erbb2/Neu* mice and multiple human cancer cohorts in this proof-of-concept study. Specifically, we identified CMBs and CMB tumor subtypes from WSIs of treatment-naïve needle biopsies of mammary tumors in MMTV-*Erbb2/Neu* transgenic mice. These models were then applied to human cancer cohorts to assess their translatability and predictive power in clinical settings. We analyzed the association of these CMB tumor subtypes with chemotherapy responses and clinical outcomes across multiple human cohorts. We performed co-enrichment analyses and revealed significant associations between CMB tumor subtypes and vital biological pathways in mice and humans, underscoring their relevance to tumor biology, treatment response, and translatability. Our study bridges the gap between experimental research and clinical application, contributing to developing more effective and personalized treatment strategies for human cancer patients (14) and, ultimately, providing a new avenue to facilitate the translation of findings in animal studies to successful clinical trials.

## RESULTS

### Study Design and Mouse and Human Cohorts

To investigate the predictive capacity of CMBs in a controlled and genetically tractable setting, we leveraged the MMTV-Erbb2/Neu transgenic mouse model bred on a C57BL/6 × FVB F1 backcross background. This model recapitulates key molecular and pathological features of human HER2+ breast cancer, while the genetic diversity introduced by the F1Bx cross enhances inter-tumoral heterogeneity and better reflects the clinical diversity observed in patients (15). Moreover, it allows for a rigorous evaluation of treatment response and biomarker discovery under standardized preclinical conditions. Importantly, prioritizing the identification of CMBs in this cross-species framework enhances their translational potential, as biomarkers conserved from mouse to human are more likely to capture fundamental aspects of tumor biology and remain valid across distinct tumor types and patient populations, unlike cohort-specific features derived solely from human data. Given the aforementioned notes, this study aimed to develop and evaluate cross-species CMBs to facilitate the translation of drug and therapeutic responses, as well as long-term outcomes from mouse models to successful human clinical studies (**Figure 1A**) through the four steps outlined below:

**Figure 1.**
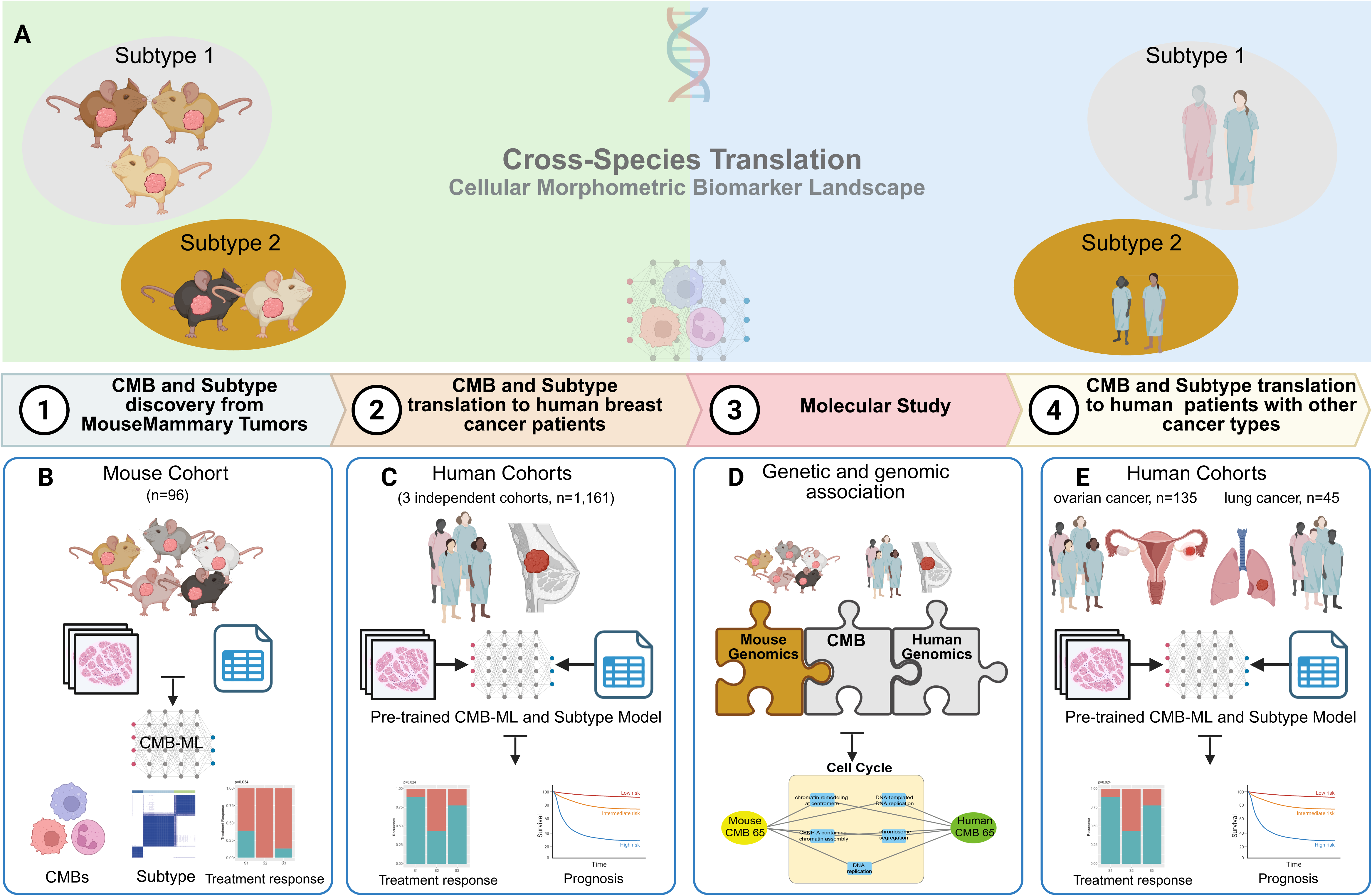
AI-Powered CMB Pipeline for Cross-Species Translatability: From Mouse Models to Human Cancer Treatment Prediction. A) Overview of the AI-driven pipeline: CMBs are extracted from whole slide images (WSIs) of mammary tumors in genetically diverse MMTV-*Erbb2/Neu* mice and applied to predict treatment response in human HER2+ cohorts. B) CMB subtype discovery in mice: Pre-treatment needle biopsies from untreated tumors were analyzed using a machine-learning pipeline (CMB-ML) to define tumor subtypes, which were retrospectively associated with chemotherapy response (docetaxel or doxorubicin). C) Translation to human breast cancer cohorts: Mouse-derived CMB subtypes were applied to three HER2+ breast cancer datasets (Spain, Yale, and TCGA-BRCA) to evaluate their predictive value for treatment response and clinical outcomes. D) Cross-species molecular characterization: Transcriptomic and genomic profiling revealed conserved biological pathways between murine and human CMB subtypes, supporting their translatability. E) Tissue-agnostic CMB application: The same mouse-derived CMB subtypes were tested in ovarian and lung cancer cohorts to explore their generalizability across different tumor types.

#### (1) Discovery of CMBs and CMB tumor subtypes from a mouse model

A genetically diverse mouse cohort was used to identify CMBs on WSIs of treatment-naïve needle biopsies of mammary tumors in 96 MMTV*-Erbb2/Neu* transgenic mice. These mice with matching H&E-stained WSIs were divided into two treatment groups: 47 were treated with docetaxel, and 49 were treated with doxorubicin (**Figure 1B; Supplementary Table S1**). After identifying the CMBs from pre-treated samples, these CMBs were utilized to identify mouse CMB tumor subtypes and evaluate their responses to treatment. Additionally, gene expression and genetic profiling of this mouse cohort were conducted to annotate the biological function of CMBs and examine the interaction between genetic variants and CMBs on treatment response.

#### (2) Translation and evaluation of CMBs and CMB tumor subtypes in human breast cancer

In addition to the mouse cohort, we analyzed three human HER2+ breast cancer (BC) cohorts (**Figure 1C**): the Spain HER2+ BC cohort (n=52), the Yale HER2+ BC cohort (n=85), and the TCGA-BRCA cohort (n=1024).

The Spain cohort included patients diagnosed at the University Hospital of Salamanca, with diagnostic slides and clinical data used to map mouse-derived CMB tumor subtypes and evaluate their association with clinical outcomes and treatment response (**Supplementary Table S2**).

The Yale cohort consisted of patients from Yale School of Medicine and was used to test the predictive value of mouse-derived CMBs and CMB tumor subtypes for trastuzumab response (**Supplementary Table S3**) (16,17).

In the TCGA-BRCA HER2-enriched cohort (n=78), we applied the CMB tumor subtype model to explore associations with recurrence status and recurrence-free survival (**Supplementary Table S4**) (18). To further assess the translational potential of CMBs beyond HER2+ BC, we also analyzed additional TCGA-BRCA subtypes, including Basal (n=181), Luminal A (n=528), Luminal B (n=200), and Normal-like (n=37) cases (**Supplementary Table S4**).

#### (3) Molecular annotation of CMBs and CMB tumor subtypes

To investigate the molecular mechanisms underlying the cross-species translatability of CMBs and CMB tumor subtypes from mouse mammary tumors to human breast cancer, we performed gene expression profiling in both our mouse cohort and used the gene expression data from the TCGA-BRCA dataset. Using the CMB model developed in mice, we built a cross-species co-enrichment network to identify shared transcriptional programs (**Figure 1D**).

#### (4) Translation and evaluation of CMBs and CMB tumor subtypes in additional human cancers

Beyond the human BC cohorts, we included one high-grade serous ovarian cancer (HGSOC) cohort (n=158) (19,20), and one lung squamous cell carcinoma (TCGA-LUSC) cohort (n=45) (**Supplementary Tables S5, S6**).

HGSOC biopsies were obtained at the time of primary debulking surgery, before chemotherapy. After excluding samples with mixed primary/metastatic origin (n=10) and those from patients who received neoadjuvant therapy (n=13), the cohort included a balanced number of chemo-refractory (n=83) and chemo-sensitive (n=52) tumors. Refractory tumors were defined as those showing progression or stable disease within six cycles of initial platinum/taxane therapy. Sensitive tumors responded to the same regimen and showed no progression within two years (**Supplementary Table S5**).

The TCGA-LUSC cohort included patients treated with platinum/taxane therapy, selected from cBioPortal based on the availability of diagnostic slides (**Supplementary Table S6**).

In summary, we integrated data from mouse models and multiple human cancer cohorts to evaluate the cross-species translatability of CMBs and CMB tumor subtypes across tumor types and treatment contexts, aiming to uncover potential mechanistic insights (**Figure 1**).

### Identification of CMB Tumor Subtypes and Their Association with Treatment Response

To evaluate the predictive potential of CMBs for treatment response in HER2+ BC, first we performed an in-depth analysis using the MMTV-*Erbb2/Neu* transgenic mouse model to identify the CMB. Ninety-six mice from a C57/FVB F1 backcross were randomized into two treatment groups: 47 treated with docetaxel, and 49 treated with doxorubicin (**Supplementary Table S1**). Needle biopsies were obtained before treatment for pathological and transcriptomic analyses.

The CMB-ML pipeline was optimized and trained on whole-slide images (WSI) of Hematoxylin and Eosin (H&E)-stained Formalin-Fixed Paraffin-Embedded (FFPE) sections from needle biopsies. This training set identified 256 distinct CMBs based on pre-quantified cellular objects randomly chosen from WSIs (**Figure 2A**; **Supplementary Figure S1**; **Supplementary Table S7**). These AI-learned CMBs represent distinct micro-anatomical patterns discovered *de novo*. The CMB-ML pipeline objectively quantifies hundreds of these high-dimensional morphological archetypes (predominantly reflecting diverse nuclear morphologies, see **Supplementary Figure S1**), enabling robust translation (8,9,11,13). After training, the CMB-ML model represented each cellular object as a sparse combination of the pre-identified 256 CMBs, resulting in a novel representation of each object as a 256-dimensional sparse code. This enabled the aggregation of CMB representations for each tumor sample.

**Figure 2.**
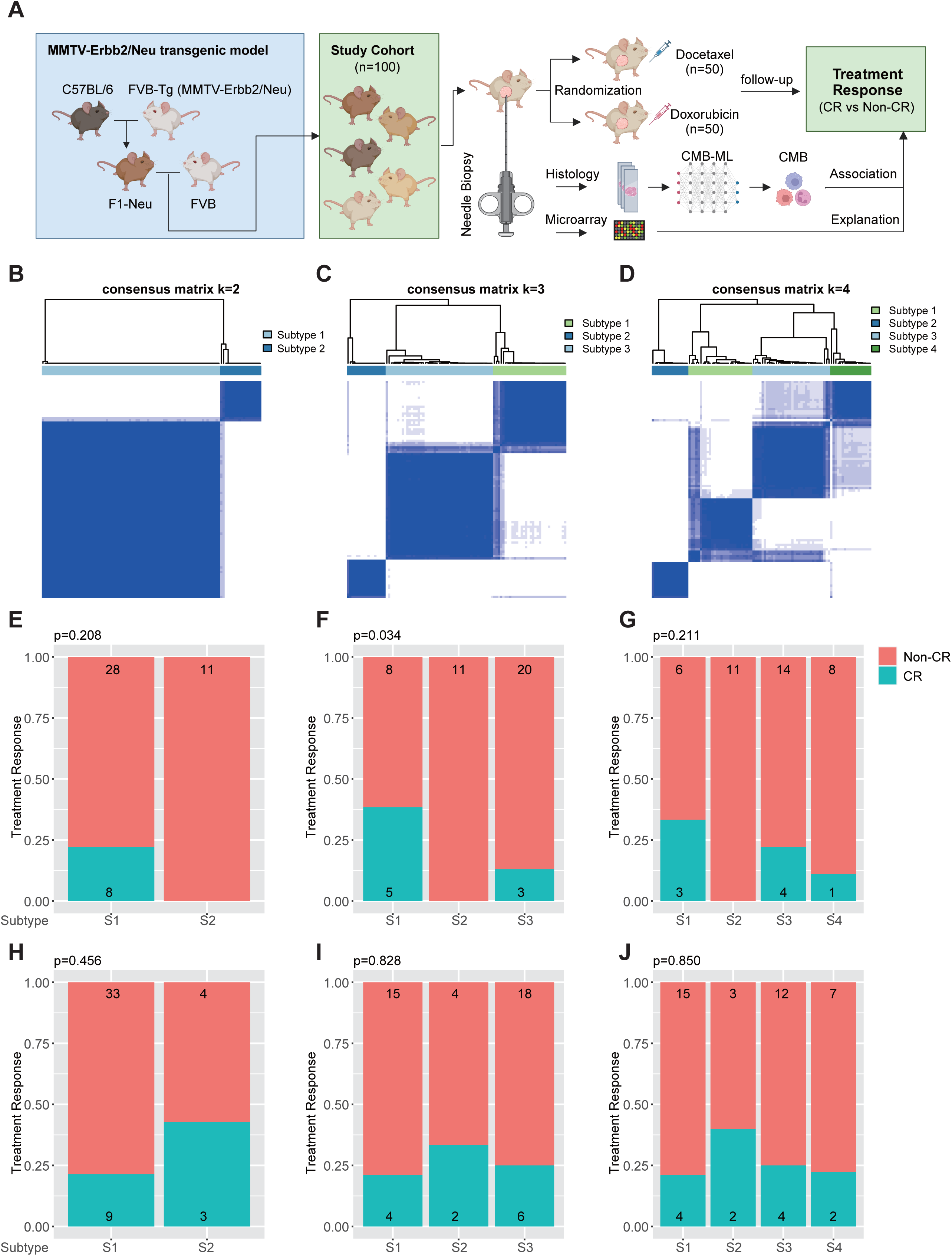
Study Design, Identification of CMB Tumor Subtypes, and Treatment Response Stratification in the Mouse Model. **A**) Schematic representation of the MMTV-*Erbb2/Neu* transgenic mouse study. Mice were randomly assigned to Docetaxel (n=47) or Doxorubicin (n=49) treatment arms. Pretreatment needle biopsies were collected for histological analysis and CMB identification using a machine learning pipeline (CMB-ML). **B–D**) Consensus clustering heatmaps showing the stability of tumor subtype classification at k = 2, 3, and 4. **E–G**) Association between CMB tumor subtypes and treatment response (CR vs. Non-CR) in mice treated with Docetaxel. Stratification was most effective at k = 3. **H–J**) Lack of significant association between CMB tumor subtypes and treatment response in mice treated with Doxorubicin across all k values. P values were calculated using the Chi-square test and are shown within the plots. Panel A was created with BioRender.com.

Consensus clustering was applied to identify the optimal number of CMB tumor subtypes for stratifying chemotherapy response. We evaluated k = 2, 3, and 4 clusters (**Figure 2B-D**). For docetaxel, k = 3 (S1, S2, S3) provided the most effective stratification, significantly distinguishing tumors based on treatment response (**Figure 2F**). In contrast, k = 2 and k = 4 did not yield meaningful differences (**Figure 2E, 2G**). Additionally, no significant associations were observed for doxorubicin across any clustering (**Figure 2H-J**).

These findings highlight the predictive value of CMBs in stratifying mouse *Erbb2/Neu+* mammary tumors and their potential for guiding personalized treatment strategies.

### Translation of mouse-derived CMBs and CMB Tumor Subtypes to the Spain HER2+ Breast Cancer

We assessed the translatability of the pre-built CMB-ML model and CMB tumor subtypes from mouse *Erbb2/Neu+* mammary tumors to a human HER2+ breast cancer (BC) cohort from Spain (**Figure 3A**; **Supplementary Table S2**). The CMB representation for each human BC sample was generated using the same strategy and pre-trained model as in the mouse cohort. Patient stratification in the Spain HER2+ BC cohort was significantly associated with treatment response over the entire follow-up period and at 3 and 5 years of follow-up (**Figure 3B-G**; **Supplementary Figure S2**), provided significant and independent prognostic value after adjusting for age and stage (**Figure 3H**), and, when combined with clinical factors, exceeded the prognostic power of clinical factors (**Figure 3I**). Recurrence status was evaluated across different CMB tumor subtype configurations, revealing consistent associations with clinical outcomes. Notably, the clinical benefit trends were highly consistent with the observations in the mouse cohort, reinforcing the robustness and translatability of CMB tumor subtypes from mice to human BC patients. Additionally, a LASSO model with bootstrapping further demonstrated the predictive power of mouse-derived CMBs in identifying recurrence risk in the Spain HER2+ BC cohort (**Supplementary Figure S3A-D**).

**Figure 3.**
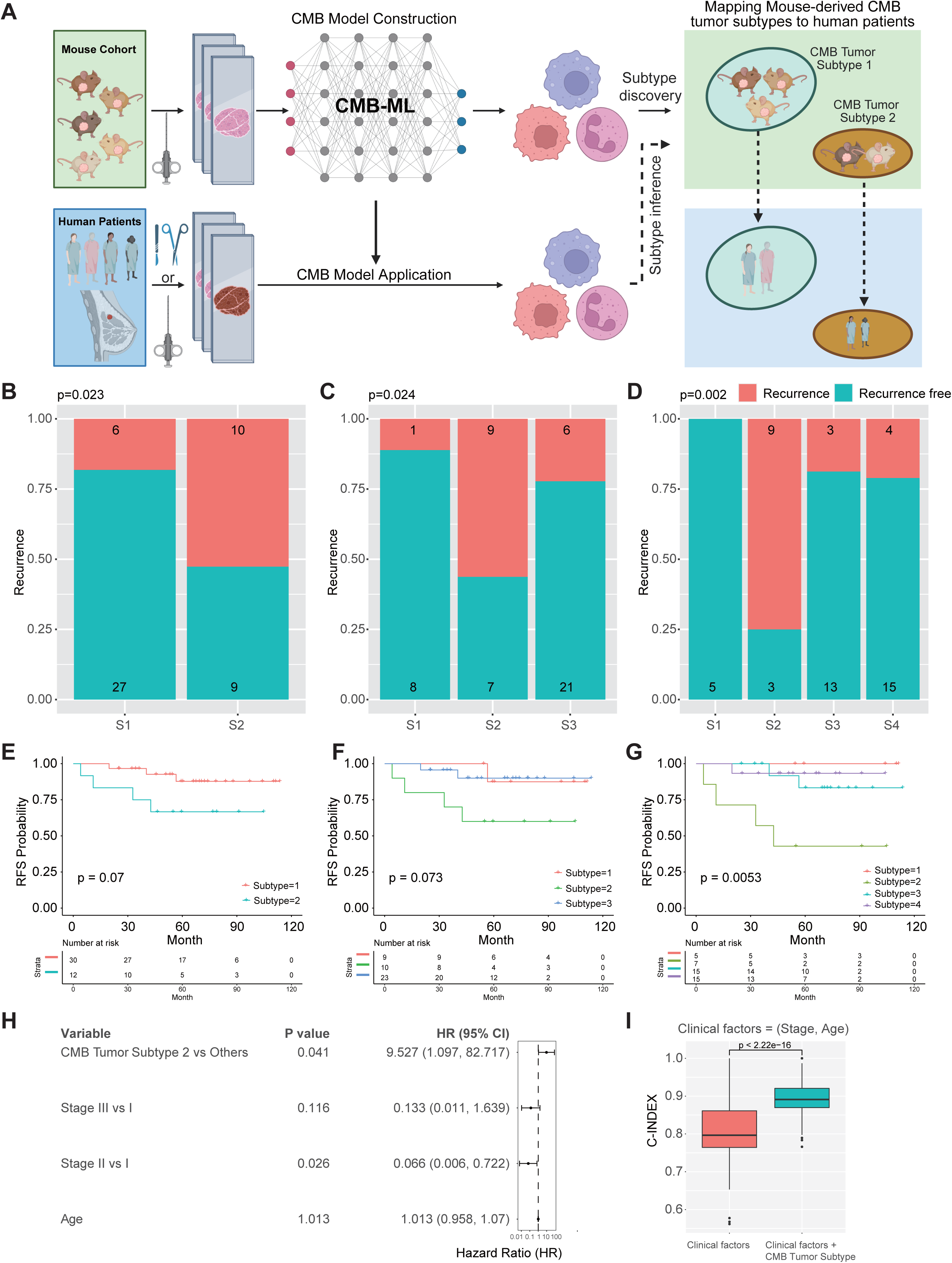
Translating Mouse-Derived CMB Subtypes to Predict Recurrence in the Spain HER2+ Breast Cancer Cohort (SP-BC-HER2). **A**) **Overview of the CMB-ML translation process:** Mouse-derived cellular morphometric biomarkers (CMBs) and their associated tumor subtypes were mapped onto human HER2+ breast cancer samples using a pre-trained CMB-ML model. The model was applied without additional retraining to test whether CMB-defined tumor subtypes maintain prognostic value in humans. **B–D**) **Recurrence status** across different CMB tumor subtypes in the SP-BC-HER2 cohort. **E–G**) **Recurrence-Free Survival (RFS) analysis** evaluating the prognostic value of CMB-defined subtypes. **H**) Multivariate Cox proportional hazards (PH) regression analysis demonstrates the independent and significant prognostic value of mouse-derived CMB tumor subtypes (k=4 model) in Spanish HER2+ breast cancer patients, after adjusting for age and stage. **I**) The multimodal integration of clinical factors and CMB tumor subtype significantly outperforms clinical factors in prognostic power, where C-Index was evaluated with a bootstrapping strategy (80% training, 20% testing, 1,000 iterations). P values for (B–D) were obtained using the Chi-square test, for (E–G) using the log-rank test, for (H) using multivariate CoxPH regression, and for (I) were acquired using the non-parametric Mann-Whitney test.

In summary, the application of the mouse-derived CMB model to the Spain HER2+ BC cohort identified tumor subtypes with additional prognostic value after adjusting for clinical factors, thus providing a clinically relevant tool for predicting treatment response and recurrence risk. These findings support the successful translation of CMB tumor subtypes from the mouse model to human patients.

### Translation of Mouse-Derived CMB Tumor Subtypes to the Yale HER2+ Breast Cancer Cohort and Their Association with Trastuzumab Treatment Response

Using the pre-built CMB-ML model derived from mouse mammary tumors, we mapped the Yale HER2+ BC cohort to evaluate its applicability in human patients and its association with neoadjuvant trastuzumab ± pertuzumab treatment response (**Supplementary Figure S4A-C**). Treatment response, obtained from pathology reports, was categorized as responders (complete pathologic response, pCR) or non-responders (residual invasive disease, lymphovascular invasion, or metastasis) (**Supplementary Table S3**).

When classified into two subtypes, the model showed limited stratification capability (**Supplementary Figure S4A**). However, the three-subtype model effectively distinguished between responders and non-responders (**Supplementary Figure S4B**), suggesting the identification of distinct biological behaviors. The four-subtype model also differentiated treatment responses (**Supplementary Figure S4C**), though the three-subtype classification provided the most robust stratification.

A LASSO model with bootstrapping confirmed the predictive power of mouse-derived CMBs for trastuzumab response in the Yale HER2+ BC cohort (**Supplementary Figure S4D-G**). This model (AUC: 0.804, 95% CI 0.505-1.000) outperformed an independent deep-learning model trained directly on this human dataset (AUC=0.68, 95% CI 0.47−0.88) (16), which showed lower predictive accuracy. Notably, its performance was comparable to that of a hybrid approach combining deep learning with expert annotations from a senior breast pathologist (AUC=0.80, 95% CI 0.69−0.88), which required manual curation (16).

In summary, the application of the mouse-derived CMB tumor subtype model to the Yale HER2+ BC cohort successfully identified tumor subtypes associated with trastuzumab treatment response. The three-subtype model demonstrated the highest stratification accuracy, highlighting its potential clinical utility for predicting patient outcomes and guiding treatment strategies.

### Mapping TCGA-BRCA HER2-enriched Breast Cancer Cohort to Mouse-Derived CMB Tumor Subtypes and Association with Clinical Outcome

Using the CMBs and CMB tumor subtypes identified from the mouse model, we mapped the TCGA-BRCA HER2-enriched BC cohort (**Supplementary Table S4**) to corresponding subtypes to evaluate their association with clinical outcomes, focusing on recurrence, where PAM50 molecular subtype was used due to its advantages over the classical definition using HER2 amplification (21).

Recurrence status (Recurrence vs. Recurrence-Free) was assessed across different CMB tumor subtypes and configurations (*k*=2,3,4) (**Figure 4A-C**). The three-subtype model (*k*=3) provided the most effective patient stratification, identifying distinct biological behaviors related to recurrence (**Figure 4B**). Kaplan-Meier analysis further confirmed its prognostic relevance, showing a significant association with recurrence-free survival (RFS) (**Figure 4D-E**).

**Figure 4.**
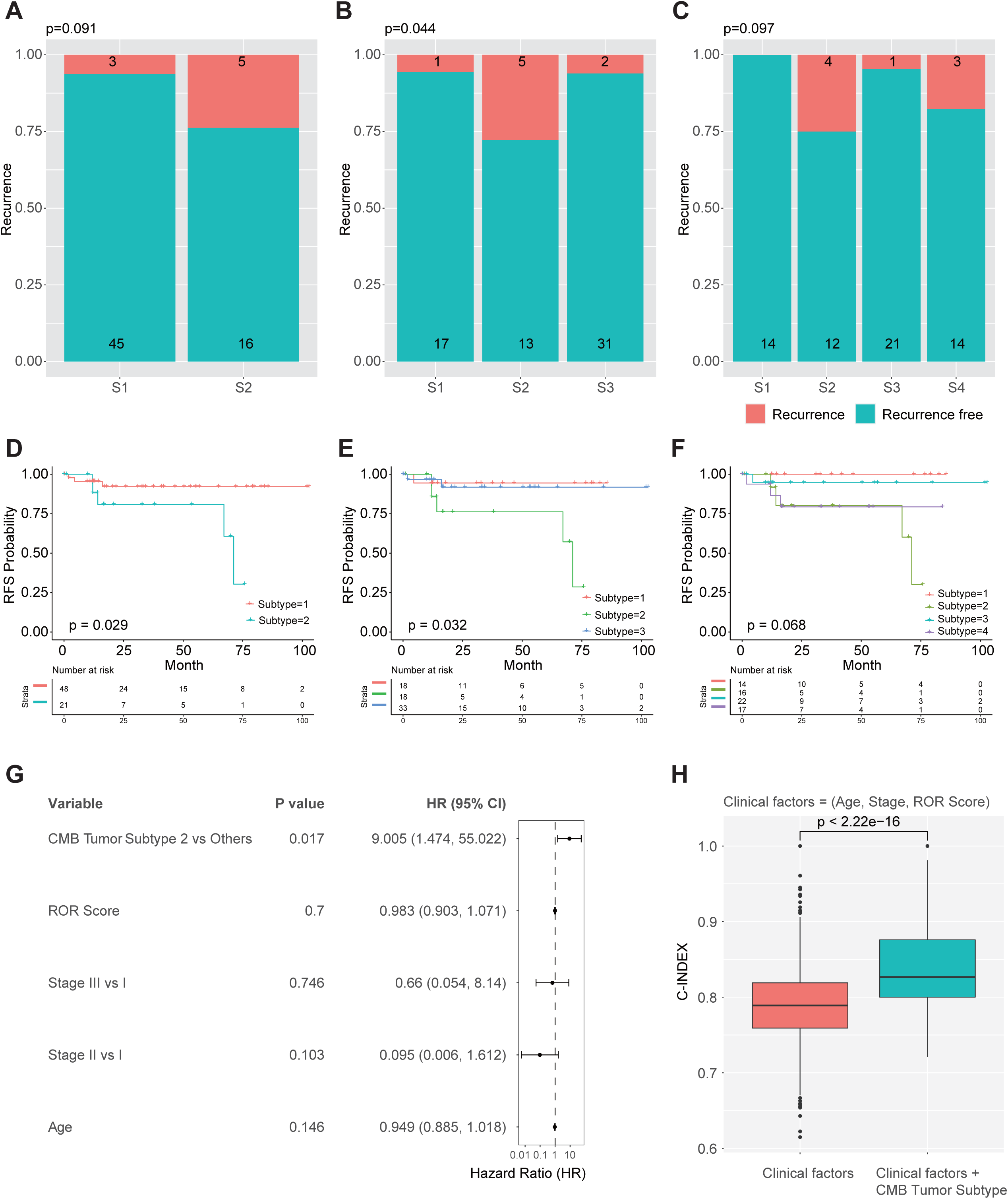
Mapping the TCGA-BRCA HER2-Enriched Breast Cancer Cohort to Mouse-Derived CMB Tumor Subtypes and Their Association with Recurrence and Survival. A–**C**) Association between CMB-defined subtypes and recurrence status in the TCGA-BRCA HER2-enriched cohort for k = 2, k = 3, and k = 4 subtype solutions. Subtype stratification is most pronounced at k = 3, while k = 2 and k = 4 show non-significant trends. **D–F**) Recurrence-free survival (RFS) analysis for the same subtype groupings, highlighting a significant survival advantage at k = 2 and k = 3, with a borderline trend at k = 4. **G**) Multivariate CoxPH regression analysis demonstrates the independent and significant prognostic value of mouse-derived CMB tumor subtypes (k=3 model) in TCGA-BRCA HER2-enriched patients, after adjusting for PAM50 ROR score, age, and stage. **H**) The multimodal integration of clinical factors and CMB tumor subtype significantly outperforms clinical factors, including PAM50 ROR score, in prognostic power, where C-Index was evaluated with a bootstrapping strategy (80% training, 20% testing, 1,000 iterations). P values for (A–C) were obtained using the Chi-square test, for (D–F) using the log-rank test, for (**G**) using multivariate CoxPH regression, and for (H) using the non-parametric Mann-Whitney test and are shown within the figure.

Importantly, the CMB tumor subtype provided additional prognostic value after adjusting for clinical and molecular factors, including the PAM50 Risk of Recurrence (ROR) score, age, and stage (**Figure 4G**). Furthermore, the combination of the CMB tumor subtype with clinical factors significantly improved the prognostic power (**Figure 4H**). In addition, the recurrence status at 3-year and 5-year time points was also assessed across different CMB tumor subtypes. Consistent but non-significant trends were observed (**Supplementary Figure S5**).

To further improve the prognostic power of mouse-derived CMBs in human breast cancer patients, we constructed the CMB risk score, which demonstrated independent prognostic value after adjusting for ROR score, age and stage, and provided distinct prognostic stratification for the entire TCGA-BRCA cohort as well as for individual PAM50 subtypes, including HER2-enriched, Basal-like, Luminal A, and Luminal B subtypes (**Supplementary Figure S6**).

In summary, applying the mouse-derived CMB-ML model to the TCGA-BRCA HER2-enriched cohort successfully identified tumor subtypes significantly associated with recurrence and RFS, and demonstrated additional prognostic value after adjusting for clinical and molecular factors. Additionally, the independent and additional prognostic value provided by the CMB risk score suggests that mouse-derived CMBs may extend beyond HER2-enriched breast cancer.

### Differentially Expressed CMB (DECMB)-Associated Molecular Co-Enrichment Network Between Mouse Mammary Tumors and Human Breast Cancer Patients

To explore the potential molecular mechanism for the translatability of CMBs/CMB tumor subtypes from mouse mammary tumors to human BC, we analyzed the differentially expressed CMB-associated co-enrichment network. The differentially expressed CMBs are defined as CMBs that are significantly different in abundance between mouse tumors in Subtype 2 and tumors in other Subtypes (1 and 3), where the subtyping model with k = 3 was selected due to its robust association with clinical outcome across species. This revealed 26 significant CMBs with 12 increased (higher abundance in Subtype 2) and 14 decreased (lower abundance in Subtype 2) (|log_2_(fold change) |>3, FDR<0.05; **Figure 5A; Supplementary Figure S1 and Supplementary Table S7**). Examining representative examples (**Supplementary Figure S1**) reveals that DECMBs enriched in poorer-responding subtypes often incorporate features suggesting nuclear atypia, while those enriched in the better-responding subtype tend to display greater regularity. This consistency with established histopathological correlates supports the biological relevance of the quantitative differences captured by the DECMBs.

**Figure 5.**
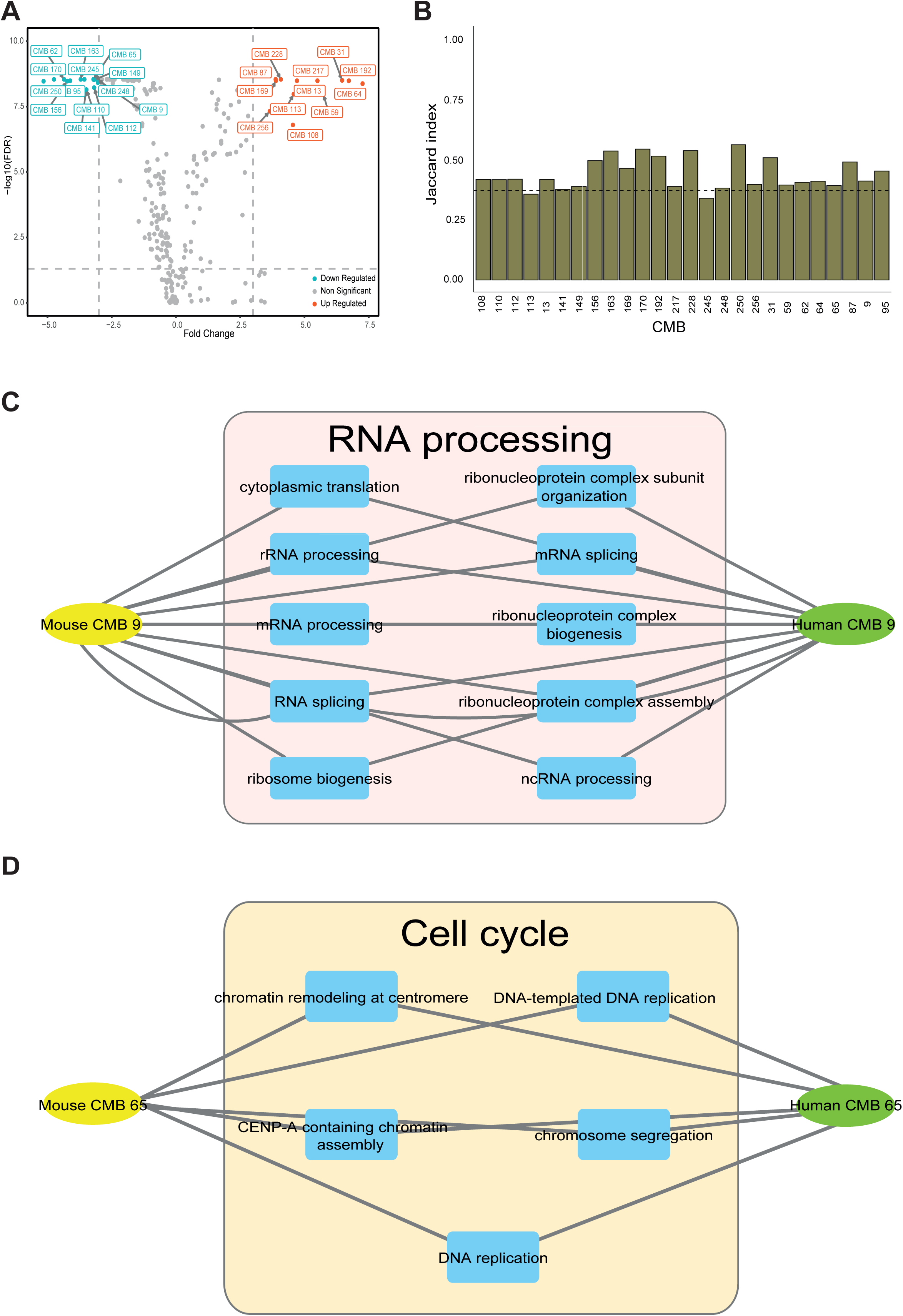
Differentially Expressed CMB (DECMB)-Associated Molecular Co-enrichment Network Between Mouse Mammary Tumors and Human Breast Cancer Patients. **A**) Volcano plot showing differentially expressed CMBs (DECMBs) between mouse tumors of subtype 2, and those of subtypes 1 and 3, with a | log_2_(fold change) | > 3.0 and false discovery rate (FDR) < 0.05. Upregulated CMBs are in red, downregulated CMBs are in blue, and non-significant CMBs are in gray. A total of 26 significant CMBs were identified, with 12 increased and 14 decreased. **B**) The association between CMBs and transcriptomics was identified in mouse mammary tumors treated with docetaxel and in human breast tumors (TCGA-BRCA HER2-enriched), followed by enrichment analysis. Jaccard index evaluation demonstrates significant conservation of 24 DECMBs between mouse and human in terms of co-enriched biological process (BP) Gene Ontology (GO) IDs, where the Jaccard indices of these 24 DECMBs exceed the 95th quantile (Jaccard index = 0.37 shown as the dash line) of the null distribution established through permutation analysis. **C**) BP enrichment analysis for CMB9 in human and mouse samples shows interconnected processes such as RNA splicing, cytoplasmic translation, and mRNA processing. **D**) BP enrichment analysis for CMB65 in both human and mouse samples, highlighting shared processes like regulation of chromosome segregation.

Next, we identified CMB-correlated genes in both mice and humans. This association was established by calculating the Spearman correlation between the abundance profile of each specific DECMB and the expression profile of each gene across all tumors within the respective cohorts (mouse or human TCGA-BRCA). Genes showing significant correlation (|R²|>0.25, p<0.05; see Methods for details) were considered CMB-correlated. Wethen compared Gene Ontology (GO) biological process (BP) terms from transcriptomic signatures associated with the 26 CMBs in both mice and humans, using the TCGA-BRCA cohort. The Jaccard index evaluation between mice and humans revealed the significantly conserved biological functions represented by BP GO IDs (details in the method section) (**Figure 5B**). For example, CMB9 shared biological processes like RNA splicing, cytoplasmic translation, and mRNA processing (**Figure 5C**); and CMB65 shared processes like regulation of chromosome segregation (**Figure 5D**). Shared biological processes between human and mouse related to some other DECMBs are shown in **Supplementary Figure S7**.

The analysis identified significant CMBs associated with chemotherapy response, with substantial overlap in biological processes between species, underscoring their translational relevance. Specific CMBs, such as CMB9 and CMB65, play conserved roles in tumor biology and may guide personalized treatment strategies in clinical settings.

### Translation of Mouse-Derived CMB Tumor Subtypes to the High-Grade Serous Ovarian Cancer (HGSOC) Cohort and Their Association with Platinum/Taxane Treatment Response

To evaluate the tissue-agnostic translatability of the pre-built CMB tumor subtype model derived from mouse mammary tumors, we applied it to the HGSOC cohort (**Supplementary Table S5**), assessing its relevance in human ovarian cancer and its association with platinum/taxane treatment response (**Figure 6A-F**). Refractory cancers were defined as those that progressed or remained stable within six cycles of initial platinum/taxane therapy after debulking surgery (*n*=83). Sensitive tumors were those that responded and remained progression-free for at least two years (*n*=52).

**Figure 6.**
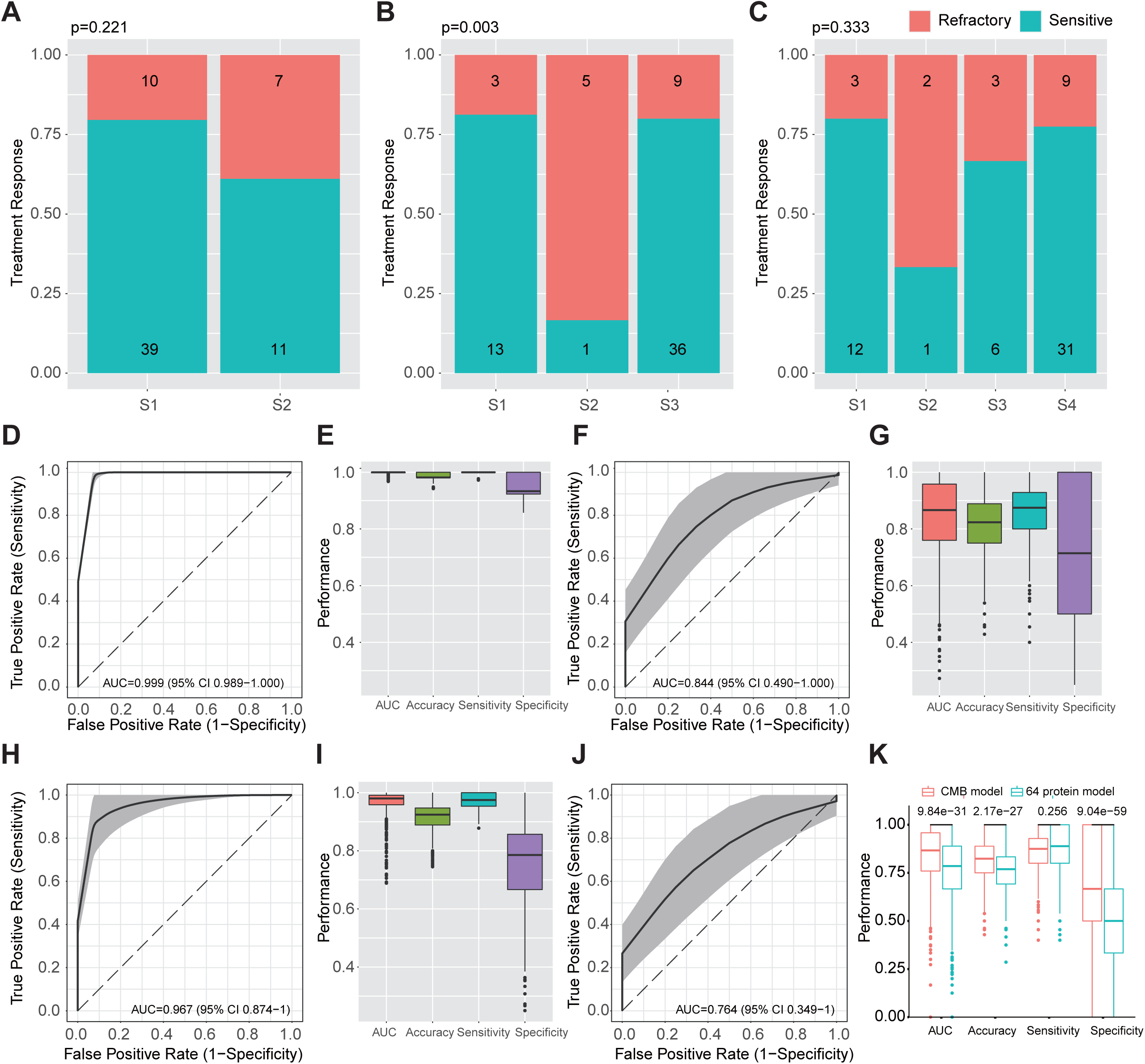
**Mapping the primary tumors in High-Grade Serous Ovarian Cancer (HGSOC) Cohort to Mouse-Derived CMB Tumor Subtypes and Predicting Platinum/Taxane Treatment Response using CMBs and proteomics. A-C**) Treatment response distribution for k = 2, k = 3, and k = 4 CMB subtypes in primary tumors of the HGSOC cohort. Only k = 3 subtypes showed significant stratification of responders and refractory tumors. **D-G**) Performance of the LASSO-based model in predicting treatment response in primary tumors using mouse-derived CMBs, evaluated in training (**G**, **H**) and testing (**I**, **J**) sets. **H-K**) Performance of the LASSO-based model in predicting treatment response in primary tumors using the 64-protein model, evaluated in training (**H**, **I**) and testing (**J**, **K**) sets, where the comparison of testing performance in (**K**) suggests that CMBs surpassed proteomics in predicting treatment response. The bootstrapping strategy (80% training, 20% testing, 1,000 iterations) was applied to assess model robustness. P values in (A-F) were calculated using Chi-square tests, and in (K) were calculated using the Mann-Whitney test.

In primary tumors, the three-subtype model (*k*=3) effectively stratified patients by treatment response, identifying distinct biological behaviors (**Figure 6B**). The two-and four-subtype models showed limited stratification (**Figure 6A, C**). In metastatic tumors, none of the models provided significant stratification (**Supplementary** Figure 6A-C).

A LASSO model with bootstrapping demonstrated accurate prediction of platinum/taxane treatment response in primary tumors (**Figure 6D-G**). Remarkably, although a prognostic classification was not achieved with the K-means clustering and chi-square method (**Supplementary** Figure 6A-C) in metastatic ovarian tumors, the LASSO model with CMBs also enabled effective prediction of treatment response in this group (**Supplementary** Figure 6D-G). Moreover, CMBs outperformed the proteomics-based models published and optimized in this HGSOC cohort (19) for predicting treatment response in both primary (**Figure 6 H-K**) and metastatic tumor stages (**Supplementary Figure S8H-K**).

In summary, the mouse-derived CMB-ML model stratified ovarian tumors by platinum/taxane response, with the three-subtype solution (k = 3) showing the strongest performance in primary tumors. Furthermore, CMBs surpassed proteomics, reinforcing their role as predictive biomarkers in ovarian cancer.

### Translation of Mouse-Derived CMB Tumor Subtypes to the Lung Squamous Cell Carcinoma (LUSC) Cohort and Their Association with Platinum/Taxane Treatment Response

To further explore the tissue-agnostic translatability of the pre-built CMB tumor subtype model derived from mouse mammary tumors, we applied it to the TCGA-LUSC cohort (**Supplementary Table S6**) to assess its relevance in human lung cancer and its association with platinum/taxane treatment response. Treatment data were retrieved from cBioPortal, and overall survival was analyzed across different CMB tumor subtype configurations (k=2, k=3, k=4).

Mouse-derived CMB tumor subtypes showed significant associations with overall survival across multiple subtype configurations (**Supplementary Figure S9A-C**) and demonstrated additional prognostic value after adjusting for age and stage (**Supplementary Figure S9D-E**), suggesting their potential clinical utility in predicting patient outcomes and guiding treatment strategies.

## DISCUSSION

The development of cancer therapies continues to rely heavily on preclinical animal models. In this proof-of-concept study, we demonstrate that cellular morphometric biomarkers (CMBs), identified through an AI-based pipeline in genetically diverse MMTV-Erbb2/Neu mice, can robustly predict treatment response and clinical outcomes in human cancer. These results highlight the potential of AI-driven, cross-species and tissue-agnostic biomarkers to enhance the translational relevance of preclinical models and support their integration into precision oncology frameworks (14).

The consistent applicability of mouse-derived CMB subtypes across multiple human datasets—including the Spain and Yale HER2+ breast cancer cohorts, the TCGA-BRCA HER2-enriched cohort (18), the HGSOC ovarian cancer cohort, and the TCGA-LUSC lung cancer cohort—demonstrates their translatability. Stratification into three CMB-defined tumor subtypes (k=3) was repeatedly associated with treatment response and prognosis, suggesting that these morphometric features capture conserved, clinically relevant biological traits that transcend tissue of origin.

Our results show that CMB-defined tumor subtypes effectively stratify HER2+ breast cancer patients by recurrence status and recurrence-free survival (RFS). The three-subtype model (k=3), which yielded the best stratification of docetaxel response in the mouse model, also translated successfully to predict trastuzumab response in the Yale HER2+ breast cancer cohort. Notably, in this cohort, the prediction model based on mouse-derived CMBs significantly outperformed a deep learning model specifically trained and optimized on the same dataset. Moreover, its performance was comparable to, and in some cases exceeded, that of a hybrid approach combining deep learning with expert annotations from a senior breast pathologist, which required manual curation (16). Beyond HER2+ breast cancer, the three-subtype model also showed strong predictive power for platinum/taxane response in both human ovarian and lung cancer cohorts. In particular, in the HGSOC cohort, the mouse-derived CMBs outperformed a 64-protein signature specifically trained within that dataset, further highlighting their cross-tumor generalizability and clinical potential.

Central to these findings is the AI-driven CMB-ML framework, which provides a new analytical lens for objective, quantitative and robust cross-species and tissue-agnostic biomarker discovery and translation. By moving beyond traditional subjective assessments, it enables the unsupervised discovery and rigorous quantification of complex, high-dimensional cellular morphometric patterns directly from H&E images. This ability to precisely measure subtle yet informative signatures was crucial for establishing robust statistical links between visual phenotypes, treatment responses, and underlying molecular programs. While the biological relevance of these AI-identified patterns is supported by their consistency with established pathological correlates, the key innovation lies in the AI’s capacity to discover and objectively measure these specific signatures. This data-driven approach effectively unlocks the rich information embedded within histology, thereby enhancing biomarker discovery and translational research.

Our study supports the conservation of cellular morphometric environments between murine models and human tumors, validating the relevance of these biomarkers in capturing key biological processes involved in cancer progression and treatment response, particularly to anthracyclines and docetaxel (1,2). The identification of shared processes—such as cell cycle regulation and chromatin remodeling—across species provides a mechanistic basis for the association between specific CMBs, tumor evolution, and therapeutic sensitivity, reinforcing their applicability in both preclinical and clinical settings (1,2). These concordant pathways suggest that the morphometric patterns quantified by the DECMBs are not merely visual correlates but surrogate readouts of underlying cell cycle and chromatin remodeling programs that modulate chemosensitivity.

Despite its promising results, this study has certain limitations. Although the MMTV-Erbb2/Neu mouse model was engineered to mimic human HER2+ breast cancer, it may not fully capture the full spectrum of molecular and phenotypic heterogeneity observed in human tumors. Moreover, the retrospective design of the analyses necessitates prospective clinical validation to confirm the predictive value and clinical applicability of the identified CMB subtypes.

Nonetheless, the use of a single, genetically diverse and well-characterized preclinical model may also be considered a major strength. The F1 backcross (F1Bx) cohort, previously shown to recapitulate substantial inter-tumoral heterogeneity observed in human breast cancer (15), provides a robust platform for biomarker discovery under standardized conditions. Importantly, identifying CMBs in a cross-species context—first in mice and then validating their conservation in humans—represents a powerful strategy to prioritize biomarkers that are more likely to be broadly applicable. Cross-species conservation increases the likelihood that these biomarkers reflect fundamental biological features and will therefore remain valid across diverse tumor subtypes and independent patient cohorts. In contrast, CMBs identified solely within a single human cohort may reflect condition-specific or cohort-specific features with limited translatability.

The consistent predictive performance of mouse-derived CMBs across multiple human cancer types, including breast, ovarian, and lung tumors, supports their cross-species and tissue-agnostic robustness and translatability. These results highlight the potential of morphometric features as fundamental, species-agnostic indicators of tumor biology and validate CMBs as cross-species biomarkers capable of predicting treatment response without relying on multiple preclinical models.

Future work should aim to extend the CMB-ML framework to additional cancer types and therapeutic contexts. Moreover, integrating CMBs with other emerging biomarkers, such as circulating tumor DNA and immune-related signatures, may offer a more comprehensive view of tumor biology and further refine patient stratification for precision oncology (5).

## Conclusion

This study demonstrates that cellular morphometric biomarkers (CMBs) identified in preclinical mouse models can be effectively translated to human cancer cohorts to predict treatment response and clinical outcomes. When integrated with molecular data, CMBs provide mechanistic insight into the cross-species conservation of tumor architecture and reveal interactions between genetic variants and morphometric features. These findings establish an AI-powered framework for cross-species translation and support the development of tissue-agnostic biomarkers for clinical use. Ultimately, this approach may enhance the translatability of preclinical models and contribute to more personalized, biomarker-guided cancer therapies. (12,14).

## METHODS

### Mice and Animal Housing

Genetically diverse F1 backcross (F1Bx) mice (n=100, **Supplementary Table S1**), generated by crossing C57BL/6J with FVB/N MMTV-Erbb2 transgenic mice, as described in our previous study (15), were treated (22) with docetaxel (n=50) or doxorubicin (n=50) in this experiment. All procedures were approved by the Institutional Animal Care and Bioethical Committee of the University of Salamanca (approval number PLE2009-0119). Details about treatment and treatment response evaluation are provided in **Supplementary Method S1**.

### RNA Extraction from Mouse Tumors

RNA extraction was performed using the Qiagen miRNeasy Mini Kit-50, which preserves miRNA populations for further analysis. The protocol followed was as previously described (15). Global RNA expression profiling was conducted using Affymetrix chips at the Genomics Unit of the Cancer Research Center, University of Salamanca.

### Gene Expression Profiling and Analysis

Details are provided in **Supplementary Method S2**.

### Human cancer patient cohorts

To evaluate the translational value of mouse-derived CMBs and CMB tumor subtypes, five human cancer cohorts were included in this study consisting of 1,341 patients from three different tumor types. Specifically, this study included the Spain HER2+ BC cohort (n=52, **Supplementary Table S2**); the Yale HER2+ BC cohort (n=85, **Supplementary Table S3**) (16,17); the TCGA-BRCA cohort (n=1,024, **Supplementary Table S4**); one human high-grade serous ovarian cancer (HGSOC) cohort (n=135, **Supplementary Table S5**); and one human lung squamous cell carcinoma (TCGA-LUSC) cohort (n=45, **Supplementary Table S6**). The human study received approval from the Ethics Board of the University Hospital of Salamanca (approval number PI 2003-03-1238). Details of these human cohorts are provided in **Supplementary Method S3**.

### PAM50 Risk of Recurrence (ROR) Score Calculation

The ROR score, derived from PAM50 subtype classification, was computed for samples in the TCGA-BRCA dataset using the ROR-S function from the genefu R package (version 2.37.0) (23) with default parameter settings. The ROR calculation methods in genefu are originally developed and described in (24), which defines a supervised risk predictor of breast cancer grounded in intrinsic molecular subtypes.

### Identification of Cellular Morphometric Biomarkers (CMBs)

We previously developed and extensively validated a powerful AI-based pipeline: Cellular Morphometric Biomarker via Machine Learning (CMB-ML), to profile cellular morphometric landscapes from WSIs in multiple types of cancers and model systems (8,9,11) that demonstrated association with prognosis, treatment response, and tumor microenvironments.

CMB-ML achieves its clinical power through the transformative quantification of tissue histology sections at the cellular level by unsupervised feature learning (25) upon fifteen critical morphometric properties, including nuclear size, chromatin density, smoothness of nuclear contour, and the background properties around nuclear regions, etc. The combinatorial quantification and sparse activation of these properties achieved by unsupervised feature learning forms the basis for each individual CMB that not only uniquely captures many tissue-agnostic concept in pathology, e.g., nuclear atypia (26–29), but also reconstructs the tissue heterogeneity, which is beyond the capability of conventional histopathological evaluation and thus provides additional clinical value to qualitative observation by human pathologists. Details of the experimental settings are provided in **Supplementary Method S4**.

### Cellular Morphometric Biomarker Profiling in Mouse Mammary Tumors and Translation to Human Breast Cancer Patients

The mouse mammary tumor subtype was identified based on mouse-level CMB representation through consensus clustering strategy (30) (ConsensusClusterPlus package in R, Version 1.50.0) with k-mean clustering, Euclidean distance and 500 bootstrapping iterations and the optimal number of subtypes was determined by the consistency of cluster assignment (consensus matrix) and the association of treatment response of subtypes. During mouse CMB tumor subtype translation, for a human breast cancer patient, the subtype was assigned as follows: (i) construct patient-level CMB representation with pre-built CMB and Subtype model; (ii) calculate the Euclidean distances between the new patient’s representation and the mean representation of each pre-identified mouse mammary tumor subtype; and (iii) assign the new patient to its closest subtype yielding smallest Euclidean distance.

### Evaluation of CMBs Association with Chemotherapy Response

Details are provided in **Supplementary Method S5**.

### Construction of CMB Risk Score and CMB Risk Stratification in TCGA-BRCA cohort

Details are provided in **Supplementary Method S6**.

### Differentially expressed CMB (DECMB) analysis in mouse cohort

DECMBs in the mouse cohort were defined as those showing significant expression differences between Subtype 2 (S2) and the other subtypes (S1 and S3), based on a Mann-Whitney non-parametric test (FDR < 0.05; |log₂FoldChange| > 3). This classification was motivated by the markedly different treatment responses observed between S2 and S1/S3.

### DECMB-associated molecular co-enrichment networks between mouse mammary tumors and human breast cancer patients

Details are provided in **Supplementary Method S7**.

## Statistical analysis

Survival differences between CMB tumor subtypes or groups were examined using the log-rank test. Differences in CMB abundance between subtypes were analyzed using the Mann-Whitney non-parametric test. Differences in the treatment response between groups were examined using the Chi-square test. P-value (FDR corrected if applicable) less than 0.05 was considered to be statistically significant. All analysis was performed with R (Version 4.0.4).

## STATEMENTS SECTION

### Data Availability

Whole slide images of the TCGA-BRCA and TCGA-LUSC cohorts were downloaded from the TCGA-GDC portal (https://portal.gdc.cancer.gov/). Clinical and molecular data were downloaded from cBioportal (https://www.cbioportal.org/). Whole slide images and clinical data of the Yale cohort (16) were downloaded from the Cancer Imaging Archive (https://doi.org/10.7937/E65C-AM96). Whole slide images and clinical data of the HGSOC cohort (20) were downloaded from the Cancer Imaging Archive (https://doi.org/10.7937/6RDA-P940). Raw data related to Spaińs mouse mammary tumor cohort and human breast cancer cohort are available upon reasonable request to corresponding authors upon IRB approval at the participating institution(s).

## Author Contributions

JPL, JHM, and HC conceptualized and designed the study. MJP and MAT performed the formal analysis. YFW, YHZ, KYJ, MHM, MME, NGS, EF, RCC, HC, JHM, JPL, NLB, and AB were involved in the critical review of the data and/or interpretation of results. MJP, HC, and JHM created the manuscript figures and supplementary materials. JPL, JHM, and HC drafted the manuscript. All authors edited, reviewed, revised, and approved the manuscript text. JPL, JHM, and HC acquired funding for the study.

## Data Availability

Whole slide images of the TCGA-BRCA and TCGA-LUSC cohorts were downloaded from the TCGA-GDC portal. Clinical and molecular data were downloaded from cBioportal. Whole slide images and clinical data of the Yale cohort were downloaded from the Cancer Imaging Archive. Whole slide images and clinical data of the HGSOC cohort were downloaded from the Cancer Imaging Archive. Raw data related to Spain's mouse mammary tumor cohort and human breast cancer cohort are available upon reasonable request to corresponding authors upon IRB approval at the participating institution(s). 

## Acknowledgment

This work was supported by the Department of Defense (DoD) BCRP, No. BC190820; and the National Cancer Institute (NCI) at the National Institutes of Health (NIH), No. R01CA184476. Lawrence Berkeley National Laboratory (LBNL) is a multi-program national laboratory operated by the University of California for the DOE under contract DE AC02-05CH11231. JPĹs lab is sponsored by Grant PID2020-118527RB-I00 funded by MCIN/AEI/10.13039/501100011039; Grant PID2023-153081OB-I00 funded by MCIN/AEI/10.13039/501100011039 the Regional Government of Castile and León (CSI144P20).

---

**Supplementary Figure S1.**
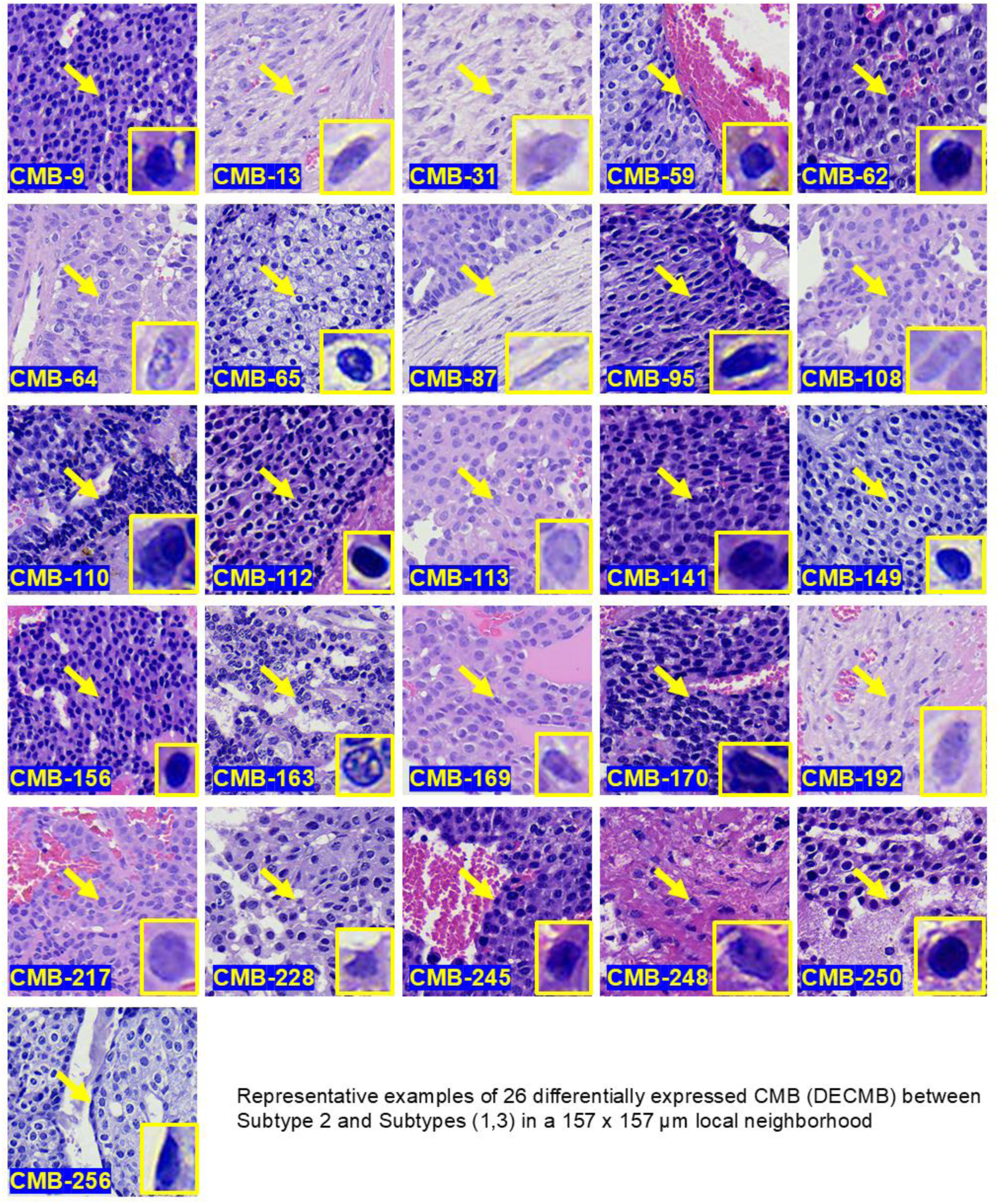
Representative examples of 26 Cellular Morphometric Biomarkers (CMBs) and the corresponding histopathological features captured by these CMBs. This panel displays representative images of the 26 CMBs extracted by the CMB-ML pipeline from pre-treatment biopsies of MMTV-Erbb2/Neu mouse mammary tumors. For each CMB, the main panel shows a 157 × 157 μm local tissue area centered on the CMB event (yellow arrow), while the inset magnifies the nuclear feature annotated. Images were uniformly contrast-enhanced for better visualization of chromatin detail without altering the original staining. These data highlight how AI-driven morphometric analysis can uncover histopathologically and clinically interpretable structures in an unsupervised manner (See Supplementary Table 8)

**Supplementary Figure S2.**
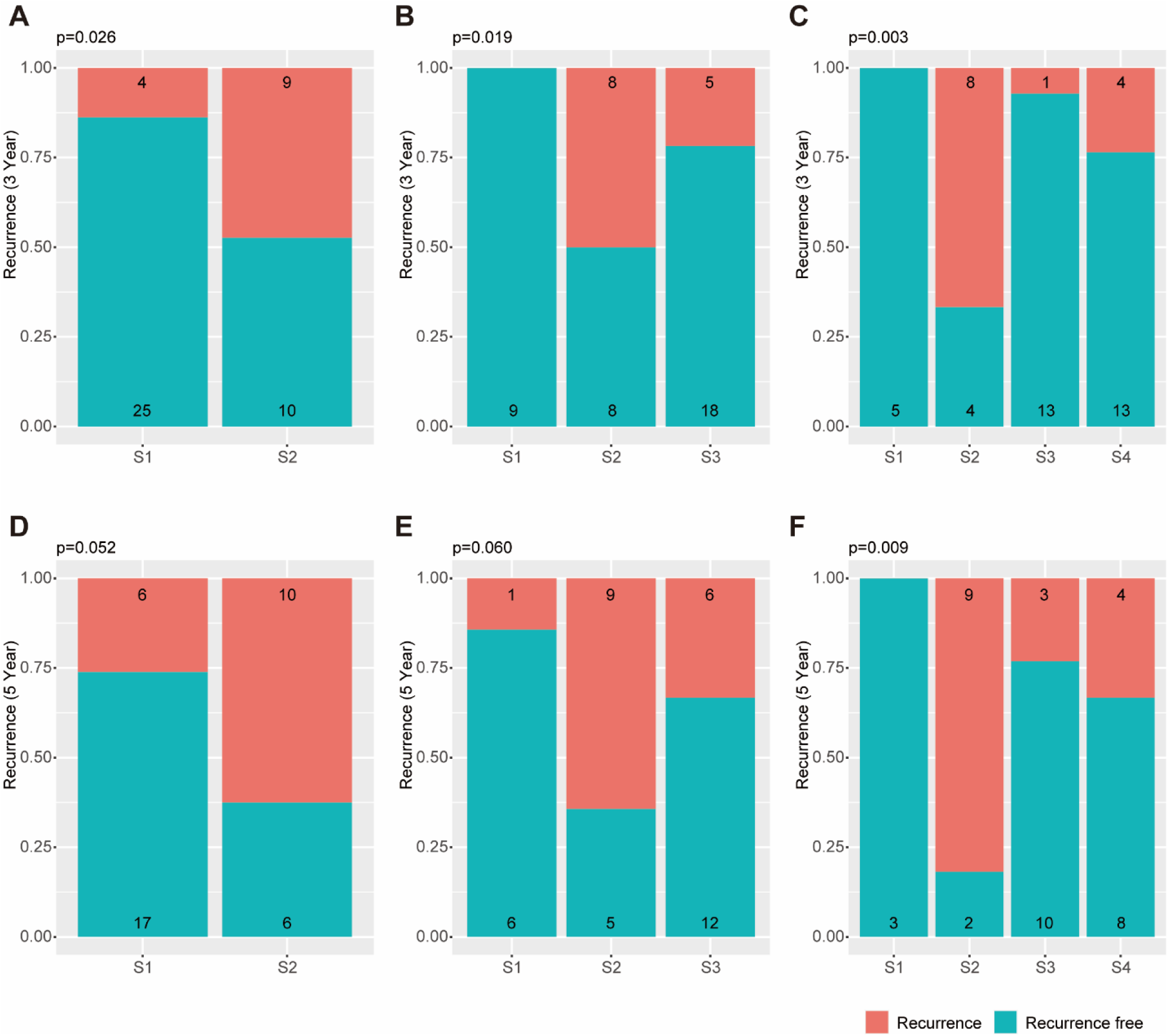
Recurrence status across CMB-defined tumor subtypes in the Spanish HER2+ breast cancer cohort (n=52), assessed at 3 years (**A–C**) and 5 years (**D–F**). Tumor subtypes were inferred using mouse-derived cellular morphometric biomarkers (CMBs) and a pre-trained CMB-ML model. Bar plots display the distribution of patients with and without recurrence, demonstrating the predictive value and translational potential of CMB-based classification. P-values were calculated using the Chi-square test.

**Supplementary Figure S3.**
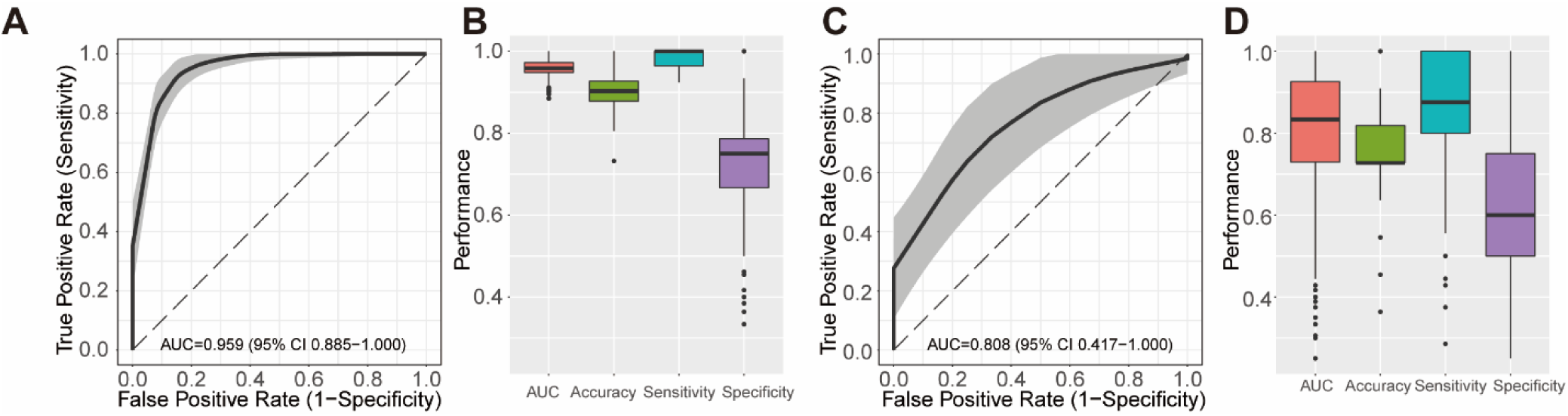
Predicting recurrence status using a machine learning approach in the Spain HER2+ Breast Cancer Cohort (SP-BC-HER2). A LASSO model with bootstrapping (80% training, 20% testing, 1,000 iterations) was used to assess the predictive performance of CMB-based classification. (A–B) Training performance was assessed using AUC, accuracy, sensitivity, and specificity metrics. (C–D) Testing performance on hold-out samples during cross-validation, demonstrating the predictive value of CMB-based classification. P values for (B, D) were obtained using the non-parametric Mann-Whitney test.

**Supplementary Figure S4.**
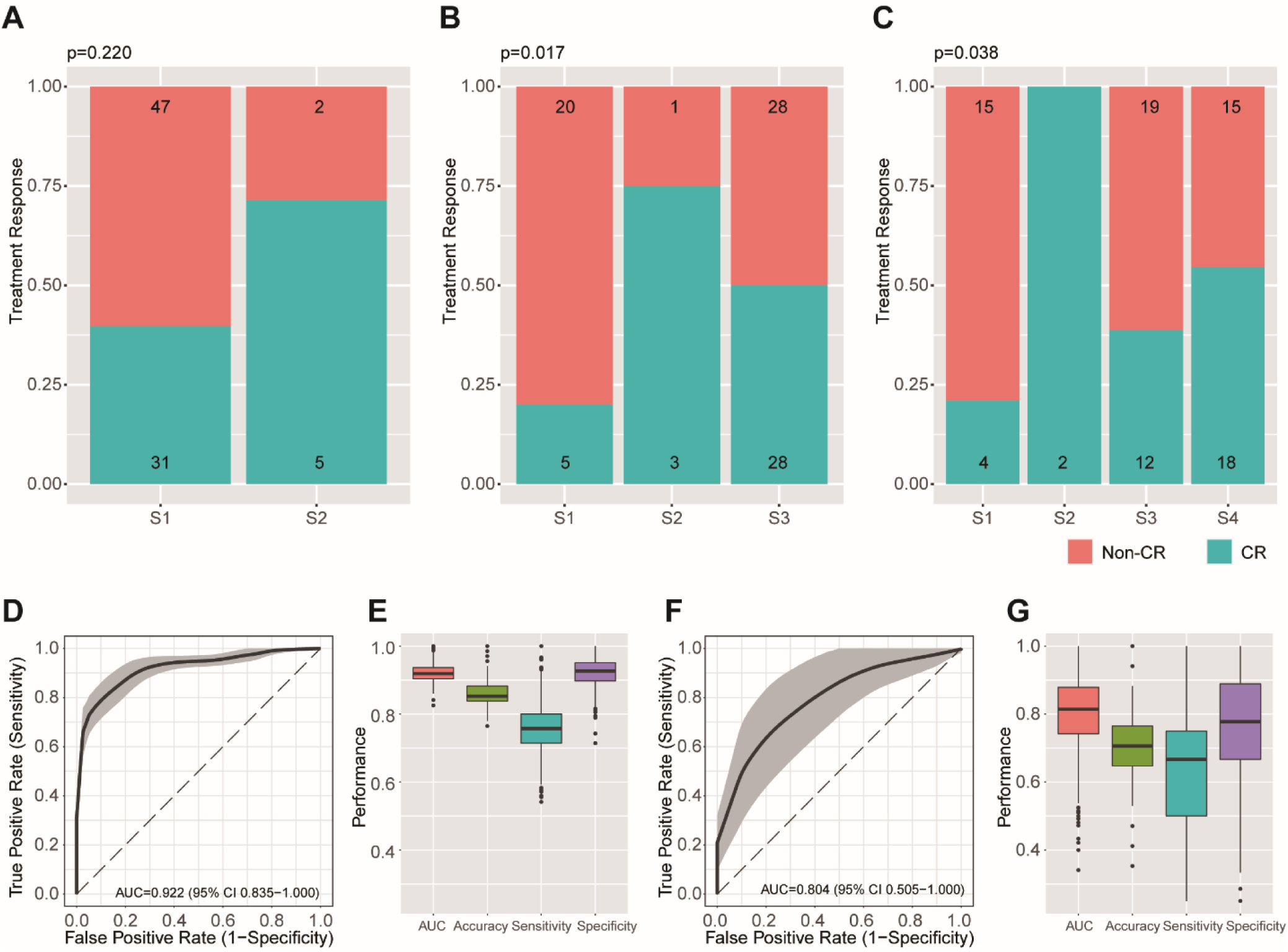
Mapping the Yale HER2+ Breast Cancer Cohort to Mouse-Derived CMB Tumor Subtypes and Predicting Trastuzumab Treatment Response. A–C) CMB-based stratification of treatment response in the Yale HER2+ cohort: Distribution of responders (CR) vs. non-responders (Non-CR) across k = 2 (A), k = 3 (B), and k = 4 (C) subtypes. Distribution of responders (CR) vs. non-responders (Non-CR) across k = 2 (A), k = 3 (B), and k = 4(C) subtypes. Improved stratification is observed at k = 3 and, to a lesser extent, at k = 4. D–G) Machine-learning prediction of trastuzumab response: A LASSO model with bootstrapping (80% training, 20% testing, 1,000 iterations) was used to predict trastuzumab response based on CMB-defined subtypes. (D–E) Training performance; (F–G) Testing performance on hold-out samples, assessed using AUC, Accuracy, Sensitivity, and Specificity metrics. P values for (A–C) were obtained using the Chi-square test.

**Supplementary Figure S5.**
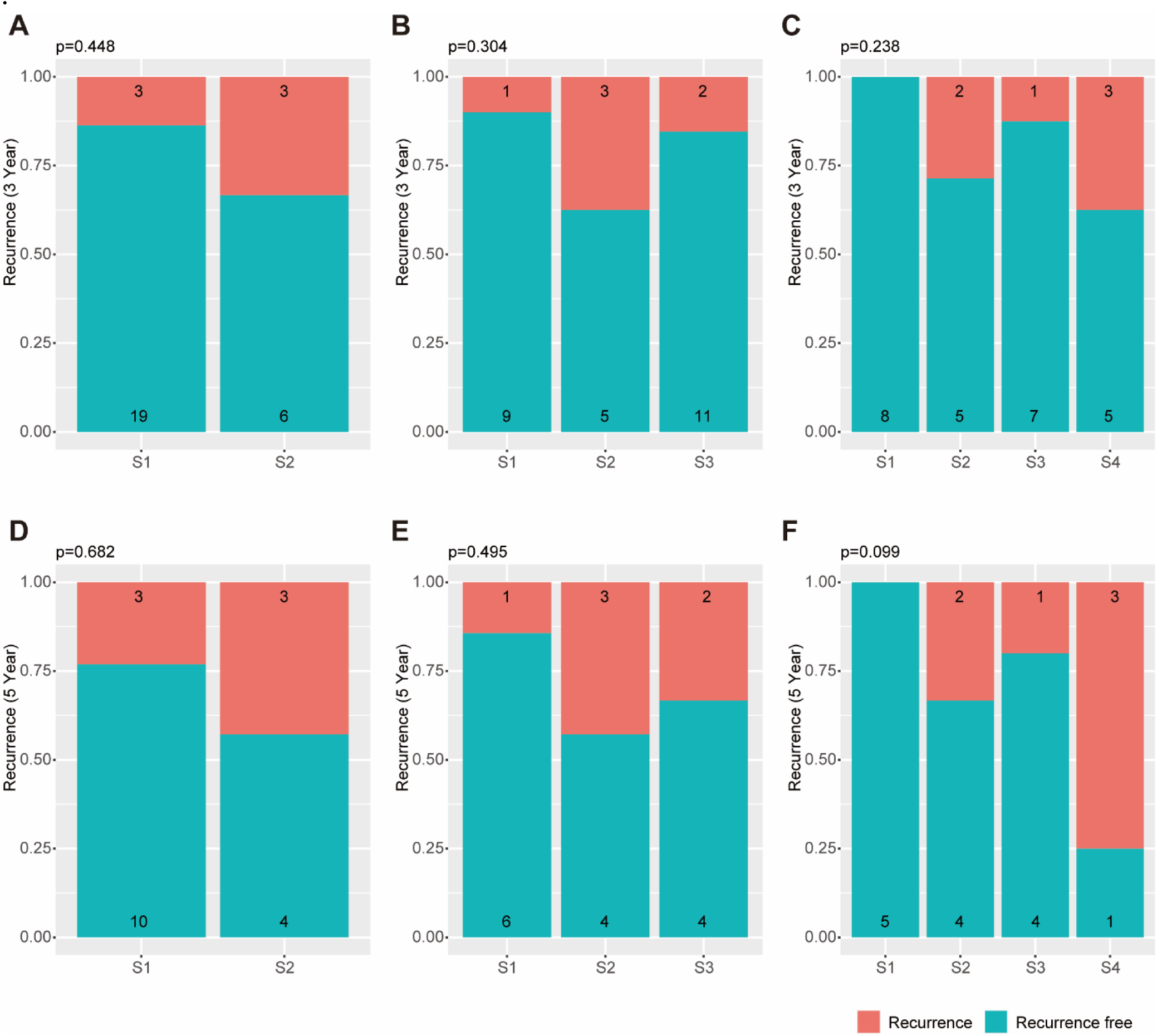
Recurrence status across CMB-defined tumor subtypes in the TCGA-BRCA PAM50 HER2-enriched breast cancer cohort, assessed at 3 years (**A–C**) and 5 years (**D–F**). P-values were calculated using the Chi-square test.

**Supplementary Figure S6.**
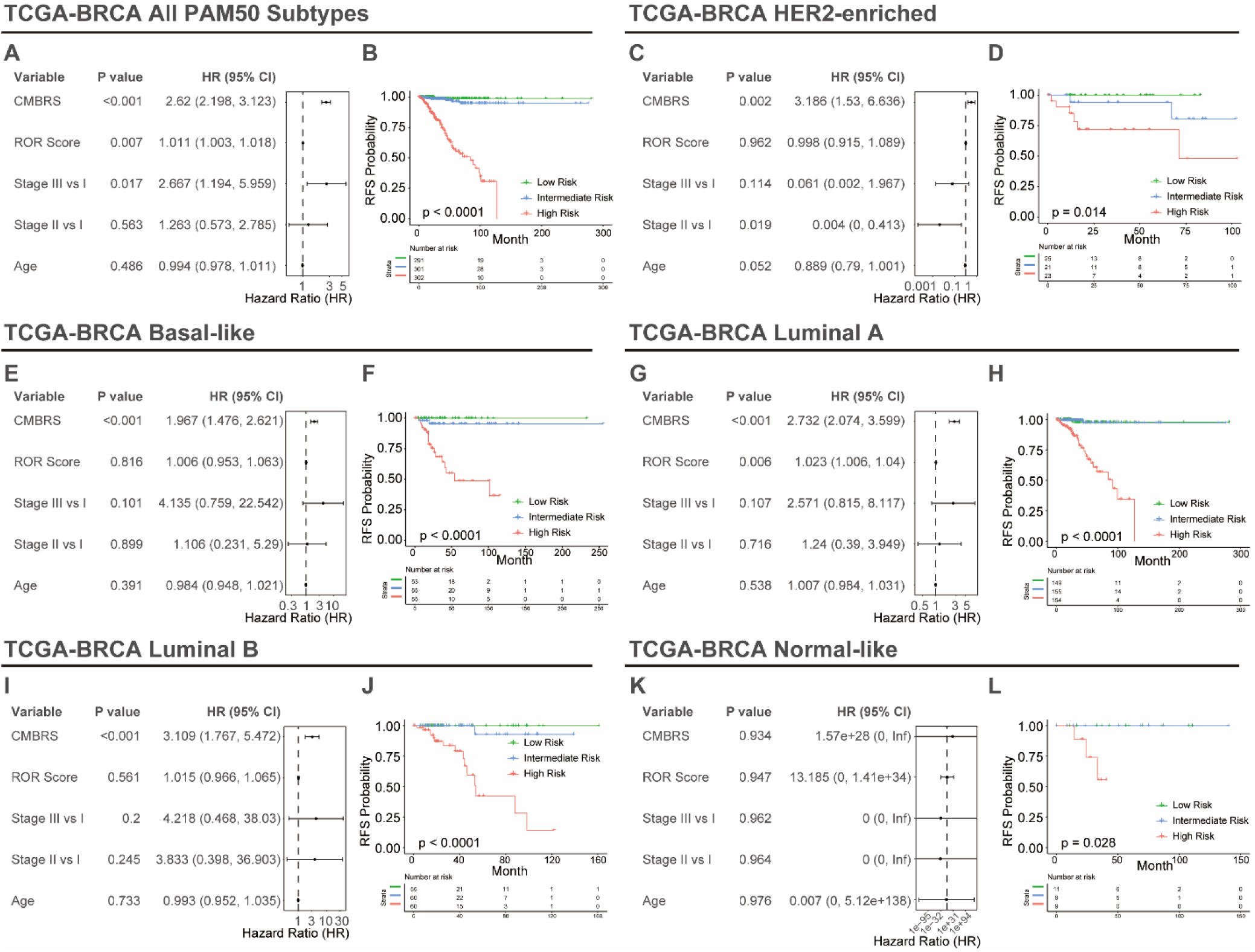
Cellular Morphometric Biomarker Risk Score (CMBRS) provides independent prognostic value (after adjusting for PAM50 ROR score, stage, and age), and stratifies the patients in the TCGA-BRCA cohort into risk groups with significantly different prognosis for combined PAM50 subtypes (A-B), HER2-enriched subtype (C-D), Basal-like subtype (E-F), Luminal A subtype (G-H), and Luminal B subtype (I-J). None of the factors provides significant prognostic value in the Normal-like subtype (K), while the CMBRS risk group still provides prognostic significance, stratification of patients in the Normal-like subtype (L). P-values in forest plots (A, C, E, G, I, K) were acquired using multivariate CoxPH regression, and in Kaplan-Meier plots (B, D, F, H, J, L) were acquired using the log-rank test.

**Supplementary Figure S7.**
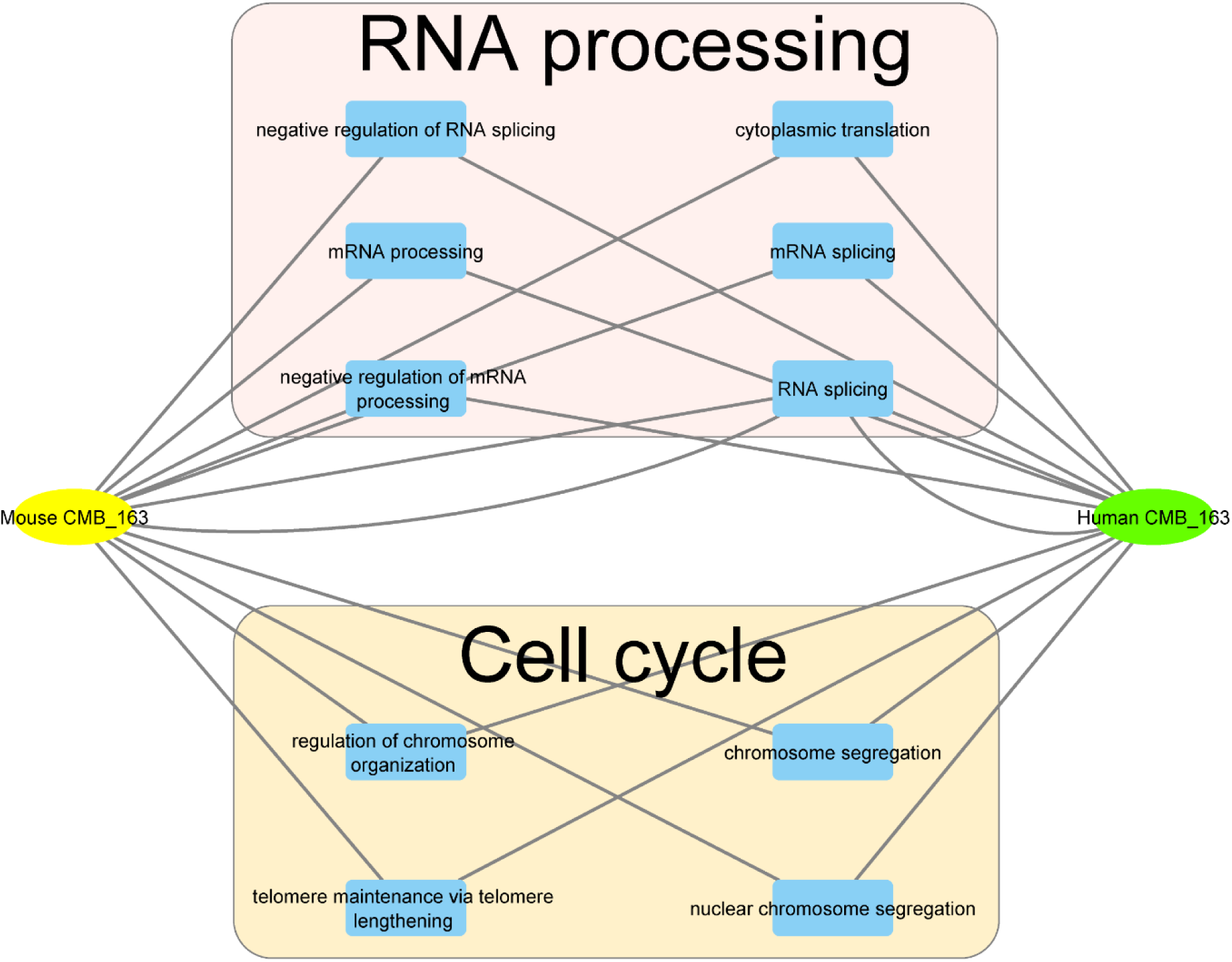
Biological processes (BP) obtained from enrichment analysis for CMB163 in human and mouse samples reveal a significant overlap in critical biological processes between the two species, highlighting the translatability of mouse-derived CMBs to human cancer studies.

**Supplementary Figure S7.**
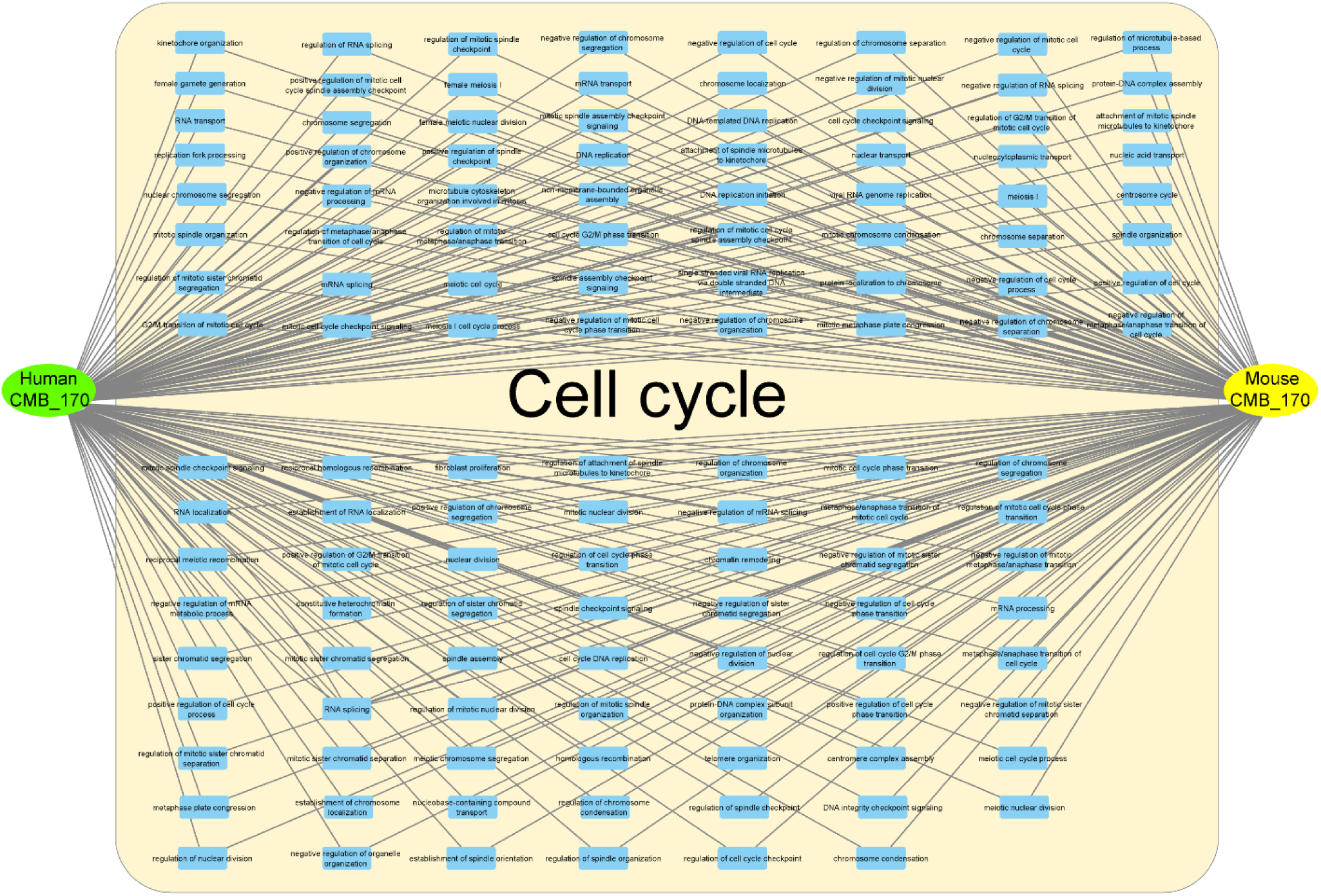
**(continued)**. Biological processes (BP) identified through enrichment analysis for CMB170 in both human and mouse samples demonstrate a significant overlap in critical biological processes between species, highlighting the translatability of mouse-derived CMBs for human cancer studies.

**Supplementary Figure S7.**
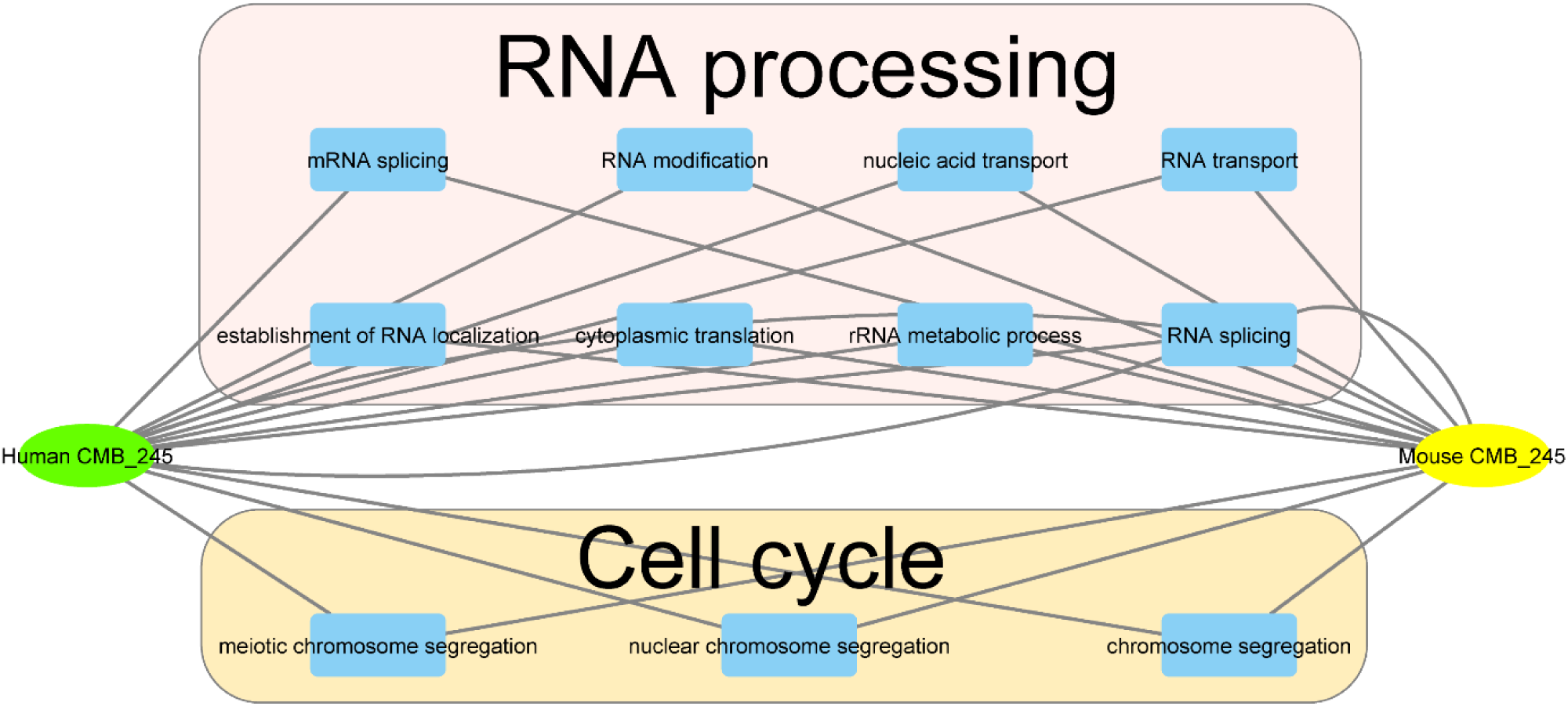
**(continued)**. Biological processes (BP) obtained from enrichment analysis for CMB245 in both human and mouse samples show a significant overlap in essential biological processes between the two species, emphasizing the translatability of mouse-derived CMBs to human cancer studies.

**Supplementary Figure S7.**
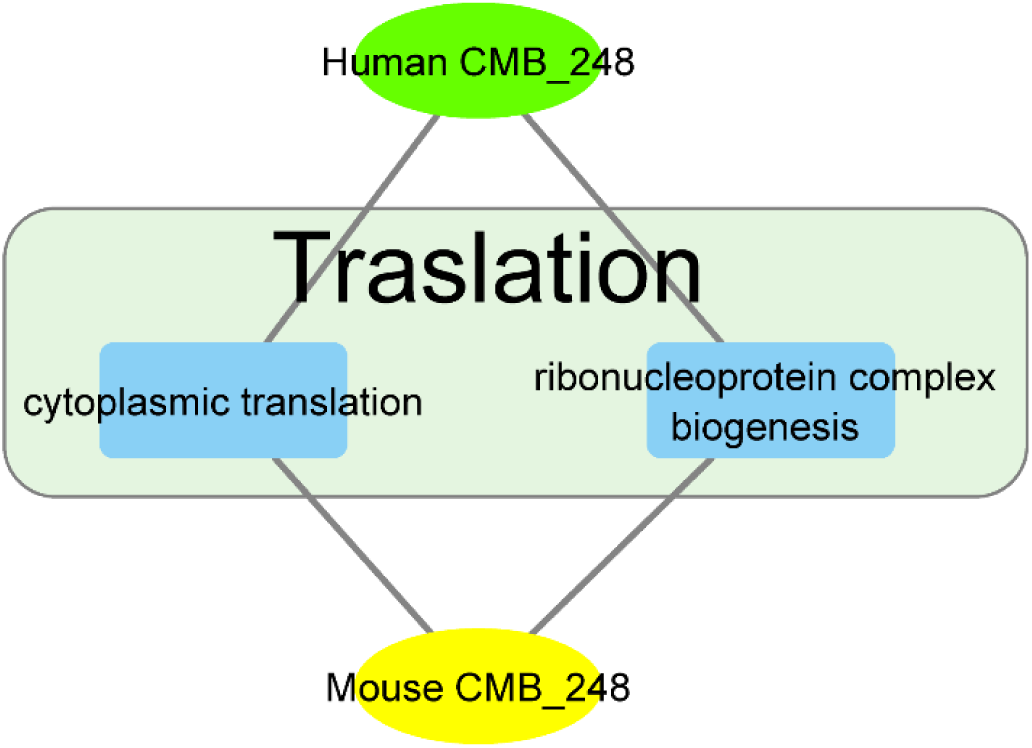
**(continued)**. Biological processes (BP) obtained through enrichment analysis for CMB248 in both human and mouse samples reveal a significant overlap in key biological processes between the species, underscoring the translatability of mouse-derived CMBs to human cancer studies.

**Supplementary Figure S7.**
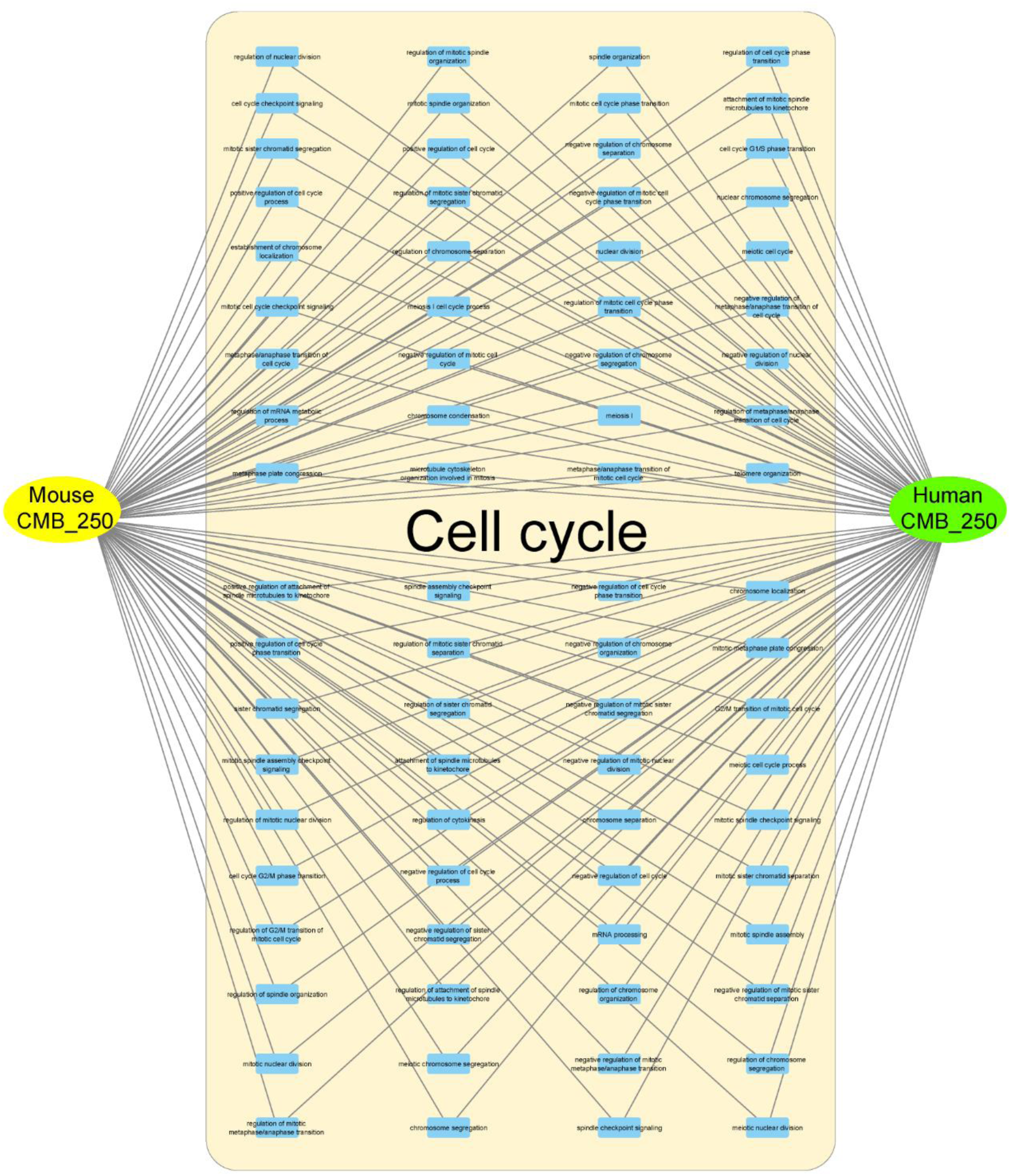
**(continued)**. The biological process (BP) derived from enrichment analysis for CMB250 in human and mouse samples shows a significant overlap in essential biological processes across species, emphasizing the translatability of mouse-derived CMBs to human cancer studies.

**Supplementary Figure S8.**
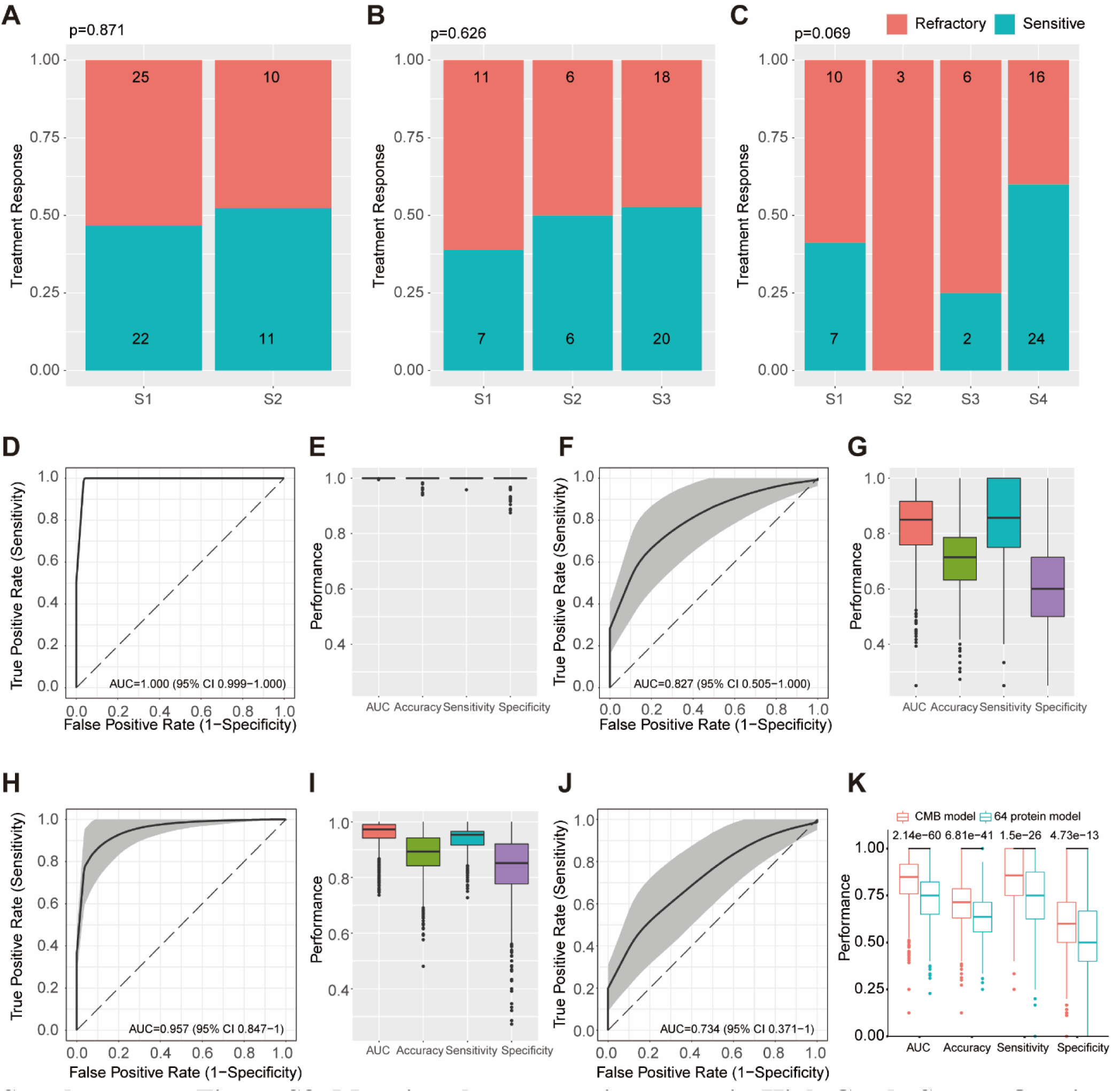
Mapping the metastatic tumors in High-Grade Serous Ovarian Cancer (HGSOC) Cohort to Mouse-Derived CMB Tumor Subtypes and Predicting Platinum/Taxane Treatment Response using CMBs and proteomics. A-C) Treatment response distribution for k = 2, k = 3, and k = 4 CMB subtypes in metastatic tumors of the HGSOC cohort. D-G) Performance of the LASSO-based model in predicting treatment response in metastatic tumors using mouse-derived CMBs, evaluated in training (G, H) and testing (I, J) sets. H-K) Performance of the LASSO-based model in predicting treatment response in metastatic tumors using the 64-protein model, evaluated in training (H, I) and testing (J, K) sets, where the comparison of testing performance in (K) suggests that CMBs surpassed proteomics in predicting treatment response. The bootstrapping strategy (80% training, 20% testing, 1,000 iterations) was applied to assess model robustness. P values in (A-F) were calculated using Chi-square tests, and in (K) were calculated using the Mann-Whitney test.

**Supplementary Figure S9.**
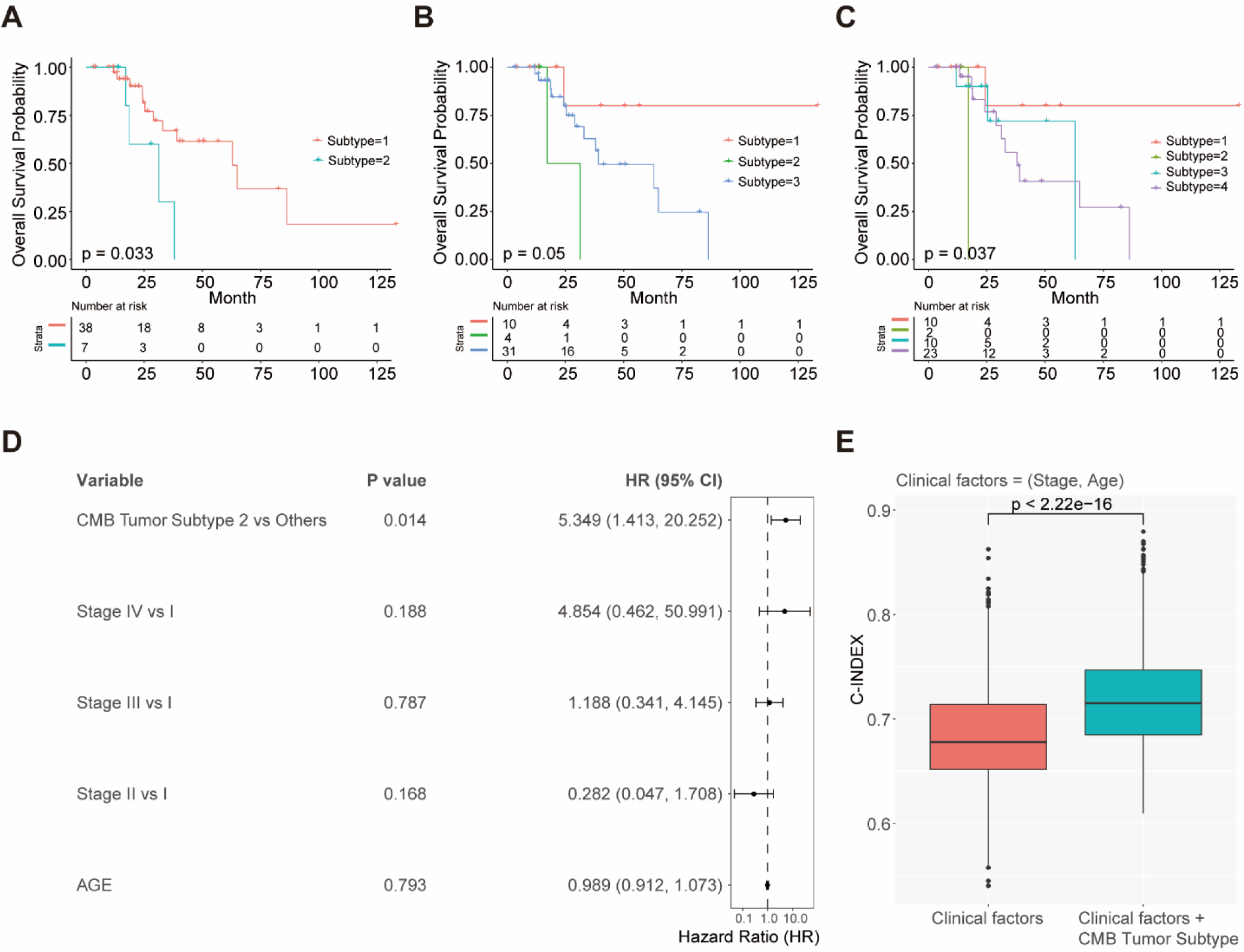
**Prognostic evaluation of mouse-derived CMB tumor subtypes in the TCGA-LUSC cohort: A-C**) Overall survival with CMB-defined tumor subtype configuration: k = 2, k = 3, and k = 4, respectively, in the TCGA-LUSC cohort, where patients were treated with platinum/taxane. **D**) Multivariate CoxPH regression analysis demonstrates the independent and significant prognostic value of mouse-derived CMB tumor subtypes (k=2 model) in TCGA-LUSC patients, after adjusting for age and stage. **E**) The multimodal integration of clinical factors and CMB tumor subtype significantly outperforms clinical factors in prognostic power, where C-Index was evaluated with a bootstrapping strategy (80% training, 20% testing, 1,000 iterations). P-values in (A-C) were calculated using log-rank tests, in (D) using multivariate CoxPH regression, and in (E) using the non-parametric Mann-Whitney test.

